# Genetic meta-analysis of levodopa induced dyskinesia in Parkinson’s disease

**DOI:** 10.1101/2023.05.24.23290362

**Authors:** Alejandro Martinez-Carrasco, Raquel Real, Michael Lawton, Hirotaka Iwaki, Manuela M. X. Tan, Lesley Wu, Nigel M. Williams, Camille Carroll, Michele T.M. Hu, Donald G. Grosset, John Hardy, Mina Ryten, Tom Foltynie, Yoav Ben-Shlomo, Maryam Shoai, Huw R. Morris

**Affiliations:** Department of Clinical and Movement Neurosciences, UCL Queen Square Institute of Neurology, University College London, UK; UCL Movement Disorders Centre, University College London, London, UK; Aligning Science Across Parkinson’s (ASAP) Collaborative Research Network, Chevy Chase, MD, 20815; Population Health Sciences, Bristol Medical School, University of Bristol, Bristol, UK; Center for Alzheimer’s and Related Dementias (CARD), National Institute on Aging and National Institute of Neurological Disorders and Stroke, National Institutes of Health, Bethesda, MD, USA; Data Tecnica International, Glen Echo, Maryland, USA; Department of Neurology, Oslo University Hospital, Oslo, Norway; Institute of Psychological Medicine and Clinical Neurosciences, MRC Centre for Neuropsychiatric Genetics and Genomics, Cardiff University, Cardiff, UK; Faculty of Health, University of Plymouth, Plymouth, UK; Translational and Clinical Research Institute, Newcastle University, Newcastle, UK; Nuffield Department of Clinical Neurosciences, Division of Clinical Neurology, University of Oxford, Oxford, UK; Oxford Parkinson’s Disease Centre, University of Oxford, Oxford, UK; School of Neuroscience and Psychology, University of Glasgow, Glasgow, UK; Department of Neurodegenerative Diseases, UCL Queen Square Institute of Neurology, University College London, London, UK; UK Dementia Research Institute, University College London, London, UK; Reta Lila Weston Institute, UCL Queen Square Institute of Neurology, London, UK; National Institute for Health Research (NIHR) University College London Hospitals Biomedical Research Centre, London, UK; Institute for Advanced Study, The Hong Kong University of Science and Technology, Hong Kong SAR, China; Genetics and Genomic Medicine, UCL Great Ormond Street Institute of Child Health, University College London, London, UK; NIHR Great Ormond Street Hospital Biomedical Research Centre, University College London, London, UK

**Author notes:** Alejandro Martinez-Carrasco; Raquel Real; Michael Lawton; Hirotaka Iwaki; Manuela Tan; Lesley Wu; Nigel Williams; Camille Carroll; Michelle Hu; Donald Grosset; John Hardy; Mina Ryten; Tom Foltynie; Yoav Ben shlomo; Maryam Shoai; Huw Morris. Corresponding authors Alejandro Martinez-Carrasco, Department of Clinical and Movement Neurosciences, UCL Queen Square Institute of Neurology, Queen Square House, Queen Square, London WC1N 3BG, England. Huw R. Morris, Department of Clinical and Movement Neuroscience, Institute of Neurology Upper 3rd Floor, Royal Free Hospital London NW3 2PF, England.

## Abstract

**Importance:** Forty percent of Parkinson’s disease patients develop levodopa-induced-dyskinesia (LiD) within 4 years of starting levodopa. The genetic basis of LiD remains poorly understood, and there have been few well powered studies.

**Objective:** To discover common genetic variants in the PD population that increase the probability of developing LiD.

**Design, setting and Participants:** We performed survival analyses to study the development of LiD in 5 separate longitudinal cohorts. We performed a meta-analysis to combine the results of genetic association from each study based on a fixed effects model weighting the effect sizes by the inverse of their standard error. The selection criteria was specific to each cohort. We studied individuals that were genotyped from each cohort and that passed our analysis specific inclusion criteria.

**Main Outcomes and Measures:** We measured the time for PD patients on levodopa treatment to develop LiD as defined by reaching a score higher or equal than 2 from the MDS-UPDRS part IV, item 1, which is equivalent to a range of 26%-50% of the waking time with dyskinesia. We carried out a genome-wide analysis of the hazard ratio and the association of genome-wide SNPs with the probability of developing LiD using cox proportional hazard models (CPH).

**Results:** This study included 2,784 PD patients of European ancestry, of whom 14.6% developed LiD. Consistent with previous studies, we found female gender (HR = 1.35, SE = 0.11, *P* = 0.007) and younger age at onset (HR = 1.8, SE = 0.14, *P* = 2 × 10^−5^) to increase the probability of developing LiD. We identified three loci significantly associated with time-to-LiD onset. **rs72673189** on chromosome 1 (HR = 2.77, SE = 0.18, *P* = 1.53 × 10^−8^) located in the LRP8 locus, **rs189093213 on** chromosome 4 (HR = 3.06,, SE = 0.19, *P* = 2.81 × 10^−9^) in the non-coding RNA *LINC02353* locus, and **rs180924818** on chromosome 16 (HR = 3.13, SE = 0.20, *P* = 6.27 × 10^−9^) in the *XYLT1* locus. Subsequent colocalization analyses on chromosome 1 identified *DNAJB4* as a candidate gene associated with LiD through a change in gene expression. We computed a PRS based on our GWAS meta-analysis and found high accuracy to stratify between PD-LID and PD (AUC 83.9). We also performed a stepwise regression analysis for baseline features selection associated with LiD status. We found baseline anxiety status to be significantly associated with LiD (OR = 1.14, SE = 0.03, *P* = 7.4 × 10^−5^). Finally, we performed a candidate variant analysis and found that genetic variability in *ANKK1* (*rs1800497*, Beta = 0.24, SE = 0.09, *P* = 8.89 × 10^−3^) and *BDNF* (*rs6265*, Beta = 0.19, SE = 0.10, *P* = 4.95 × 10^−2^) loci were significantly associated with time to LiD in our large meta-analysis.

**Conclusion:** In this association study, we have found three novel genetic variants associated with LiD, as well as confirming reports that variability in ANKK1 and BDNF loci were significantly associated with LiD probability. A PRS nominated from our time-to-LiD meta-analysis significantly differentiated between PD-LiD and PD. In addition, we have found female gender, young PD onset and anxiety to be significantly associated with LiD.

## Introduction

Parkinson’s disease (PD) is a common neurodegenerative disorder, characterised by the loss of dopaminergic neurons in the substantia nigra pars compacta. The development of levodopa induced dyskinesia (LiD) is a major clinical problem for PD patients and multiple pharmacological and neurosurgical approaches have been developed to try to prevent, attenuate or treat LiD. Dopamine (DA) is lost from the nigrostriatal pathway, which manifests as bradykinesia, muscular rigidity, rest tremor and postural instability ^1,2^. There are several symptomatic treatments for PD motor symptoms, with the metabolic precursor of dopamine, levodopa, being the “gold standard” drug. Multiple studies have shown that levodopa improves motor function as measured by the Unified Parkinson’s Disease Rating Scale (UPDRS) or the more recent MDS-UPDRS, widely used standard clinical assessments to evaluate the motor state in PD patients ^3^. However, a more recent study comparing an early treated group against a delayed treated group showed no difference in the rate of motor progression, suggesting that levodopa itself is not disease modifying ^4^. One of the major drawbacks of long-term levodopa treatment is that many PD patients experience levodopa-related motor complications, such as wearing off, dystonia and dyskinesia ^5^.

The prevalence of LiD varies across academic- and industry-led studies, averaging at around 20-40% after four years of levodopa treatment. There are two major LiD subtypes: peak-dose dyskinesia, which occur during the therapeutic window of levodopa treatment, and diphasic dyskinesia, which present at the start and end of a dose cycle^6^.

Levodopa treatment is necessary for LiD development, but there are likely to be several other mediating factors ^6^. Based on research in animal models, it is hypothesised that pulsatile delivery of oral levodopa, presynaptic nigrostriatal degeneration and intact striatal neurons are needed for the development of LiD ^6^. Major risk factors for the development of LiD include young age at onset (AAO), female gender, low body weight, disease severity, disease duration and treatment duration (from the initiation of levodopa) as well as the total dose of levodopa ^7,8^. Disease duration and treatment duration are closely related and delayed start study designs have evaluated the effect of delaying the initiation of levodopa, showing an association between longer delay and a decreased risk of LiD ^9^. There is increasing evidence to suggest that genetics plays a role in the susceptibility to LiD. Rare variants in genes such as *PRKN*, *PINK1*, and *DJ-1* have been reported to be associated with higher rates of dyskinesia ^10–12^, although patients with autosomal recessive PD usually have early onset disease, which is in itself a risk factor for LiD. A study which corrected for age and disease duration variability did not replicate the findings of a higher LiD susceptibility among *PARK2* mutation carriers ^13^.

Common variation may also influence the risk of developing LiD. Variation at the DRD2, COMT, MAOA, BDNF, SLC6A3 and ADORA2A loci have all been reported to influence the risk of developing LiD ^14–23^.

Recently, an exome-wide association study of LiD in PD found that variants in *MAD2L2* and *MAP7* loci were associated with LiD, and replicated the association of the opioid receptor gene *OPRM1* ^24^. Due to the high heterogeneity in the genetic determinants that regulate LiD, validation in large cohorts is needed.

Here, we investigated the genetic determinants of LiD by performing a meta-analysis of genome-wide survival to LiD in five different cohorts, and assessed previously reported loci. In addition, we also performed functional genetic annotation to better understand the nominated loci. Lastly, we have investigated the predictive power of a PRS, and explored baseline clinical features that were significantly associated with the development of LiD in PD using a stepwise regression approach.

## Material and Methods

The source code with all materials and methods are available on GitHub (https://github.com/AMCalejandro/LID-CPH.git; DOI: https://doi.org/10.5281/zenodo.7802142). The README explains each step of the workflow to conduct the analysis and a link to each relevant pipeline or protocol.

### Patients data and LiD definition

We accessed clinical and genetic data from the Tracking Parkinson’s (Tracking Parkinson’s)^25^, Oxford Parkinson’s Disease Centre Discovery Cohort (OPDC)^26^, Parkinson’s Progression Markers Initiative (PPMI)^27^, Parkinson’s Disease Biomarkers Program (PDBP)^28^, and simvastatin as a neuroprotective treatment for PD trial (PD-STAT)^29^ studies (eTable 1). We carried out clinical data QC on each cohort independently (eFigure 1). Levodopa is necessary for PD patients to develop LiD^6^, therefore we excluded those who were not exposed to levodopa. In addition, we removed patients who had a disease duration at study entry of more than 10 years from disease onset, patients without longitudinal data, and those with missing genotype data.

We defined PD patients as having dyskinesia if they reached an MDS-UPDRS item 4.1 score equal to or higher than 2 which is equivalent to a range of 26%-50% of the waking time with dyskinesia.. Patients were excluded if they had dyskinesia at study entry, as time to the development of dyskinesia could not be established.

### Genotype data quality control and imputation

To perform quality control (QC) at both the sample and genotype levels, we used PLINK v1.9 (RRID:SCR_001757; https://www.cog-genomics.org/plink/1.9/) ^30^. Each quality control step and the imputation approach is summarised in eMethods.

### Whole-genome sequencing data

The PDBP and PPMI cohorts included in this study were whole-genome sequenced using Illumina HiSeq X Ten Sequencer. More information can be found in https://ida.loni.usc.edu/login.jsp. WGS data was QCed using the same pipeline as the array-based data.

### Statistical analyses

We used the R programming language to perform all the statistical analysis (R Project for Statistical Computing, RRID:SCR_001905; version 4.1.3; https://www.R-project.org/).

We studied the association between genome-wide genetic variants and time to develop dyskinesia from self-reported age at PD motor onset with Cox proportional hazard (CPH) regression models under a genetic additive model, using the ‘survival’ R package (version 3.3-1; RRID:SCR_021137; https://cran.r-project.org/web/packages/survival/survival.pdf). All tests were two-tailed. To investigate the power to detect an association under a Cox regression model with the current sample size, as well as to perform a simulation on the relationship between power and allele frequency (AF), SNP hazard ratios (HR), and sample size, we used the R package survSNP (version 0.25; https://cran.r-project.org/web/packages/survSNP/index.html).

We ran time-to-LiD GWAS in each cohort separately, adjusting by AAO (or age at diagnosis in the cohorts where AAO was not available), gender, and first 5 PCs, using as our outcome the midpoint between the visit the threshold was met and the previous time point (eMethods).

Multiple studies indicate that the risk of dyskinesia relates to disease severity. To increase the power to detect genetic associations, we explored the goodness-of-fit of the model in each cohort independently after adding the following baseline covariates, which provide surrogate measures of disease severity and dopaminergic denervation at baseline: levodopa or LEDD dose, disease duration from onset to baseline assessment and baseline motor score as measured by MDS-UPDRS part III. For each cohort, we selected the model which provided the most accurate prediction of LiD based on the Akaike Information Criteria (AIC). We used the resulting model as the main model in our analysis. We summarised the nominated set of covariates in each cohort (eTable 2). We verified that the proportional hazards assumption held true by assessing the independence between scaled Schoenfeld residuals and time through the cox.zph function from the ‘survival’ package. Schoenfeld residuals are obtained by subtracting the individuals’ covariate values at the time “t” and the corresponding risk-weighted average of covariates among all those that are at risk at the time “t”. Then, they are scaled by performing a variance-weighted transformation. A non-significant relationship between the scaled residuals and time reveals proportionality of the hazards in the model.

We used METAL software (version released on the 2011-03-25; RRID:SCR_002013; https://genome.sph.umich.edu/wiki/METAL_Documentation) for meta-analysis of genome wide association summary statistics, with a fixed effects model weighted by β coefficients and the inverse of the standard errors ^31,32^. We applied a genomic control correction to the cohort-specific summary statistics by computing the inflation of the test statistic, and then applying the genomic control correction to the standard errors. We chose a meta-analysis over a merged analysis because of the heterogeneity in the inclusion and exclusion criteria across the clinical cohorts, as well as differences in the genotyping approaches (eTable 1). Statistical significance was assessed at the conservative threshold of *P* = 5 × 10^−8^, derived from a Bonferroni correction accounting for the number of independent tests and the linkage disequilibrium (LD) structure of the genome^33^.

We proved that the model met the proportional hazard assumption after including significant SNPs using the cox.zph function from the ‘survival’ package. We evaluated whether signals were replicated across different cohorts with the R package ‘forestplot’ (version 2.0.1; https://CRAN.R-project.org/package=forestplot).

### Sensitivity analyses

To validate the genome wide significance findings, we performed four sensitivity analysis to discard the associations we found in our analysis were confounded (eMethods).

### Post-GWAS analyses

We used the ‘echolocatoR’ R package (v 0.2.2; https://github.com/RajLabMSSM/echolocatoR) as a wrapper to perform fine-mapping which allows us to nominate causal variants for further study. In particular, we used the ABF approach through the ‘coloc’ R package, FINEMAP software in Unix (v v1.3; http://www.christianbenner.com/), the ‘susieR’ R package (v 0.11.92; https://cran.r-project.org/web/packages/susieR/index.html), and Polyfun-SuSiE (V1.0; https://github.com/omerwe/polyfun) ^34–38^. We produced the 95% Probability Credible Set (CS_95%_), which is the minimum set of SNPs that contains all causal SNPs with 95% probability. We reported the consensus SNPs at each locus, i.e. those that were included in the 95 CS_95%_ of at least two fine-mapping tools, therefore increasing the confidence in the nominated causal SNPs. We reported the Posterior Probability (PP) as the mean PP across all fine-mapping tools. To account for SNP LD at each region, we used the precomputed LD matrix from the UK Biobank (https://alkesgroup.broadinstitute.org/UKBB_LD/)^39^.

To evaluate the potential effect of SNPs on candidate loci on the control of gene expression we also used echolocatoR as a wrapper to access brain cell type-specific epigenetic marks from Nott and colleagues^40,41^(Data accessed using echolocatoR v 0.2.2). We mapped each locus to cell type-specific chromatin immunoprecipitation sequencing (ChIP-seq) results generated by quantifying H3K4me3 and H3K27ac epigenetic modifications, Assay for Transposase-Accessible Chromatin using sequencing (ATAC-seq) results, and Proximity Ligation-Assisted ChIP-Seq (PLAC-Seq) results, to detect and quantify chromatin contacts anchored at genomic regions. In addition, we also mapped such loci to cell-type specific transcription factors binding sites (TFBS) marks on Chip-seq experiments from the ENCODE project (RRID:SCR_006793; data accessed from echolocatoR R package v 0.2.2)^40,41^. This dataset contains 690 Chip-seq datasets representing 161 unique regulatory factors and spanning 91 human cell types. We used echolocatoR to query the ENCODE Uniform TFBS and retrieve the top 4 cell types with the highest probability density function for the top 5 regulatory elements.

To investigate whether there were several independently associated SNPs at each GWAS nominated locus, we performed a conditional and stepwise selection procedure with GCTA-COJO (version 1.93.0 beta for Linux; https://yanglab.westlake.edu.cn/software/gcta/#Overview)^42^. We used the Accelerating Medicines Partnership: Parkinson’s Disease (AMP-PD, v.2.5)^43^ data (*n* = 10,418) as the reference panel to estimate the correlation between SNPs. The reference sample was subjected to the same QC steps as described above, needed to get unbiased LD estimates ^44^.

We used the ‘coloc’ R package (version 5.1.0; https://cran.r-project.org/web/packages/colocr/index.html) to perform colocalization analysis between the SNPs associated with progression to LiD and SNPs defining gene expression in the region (eMethods). We used cis-eQTL data from MetaBrain cortex tissue^45^ (N = 6,601 individuals) and blood cis-eQTLs from eQTLGen (N = 31,684)^46^.

We used Functional Mapping and Annotation of Genome Wide Association Studies (FUMA) (RRID:SCR_017521; version 1.3.8; https://fuma.ctglab.nl/) to further characterise the nominated loci by querying GWAS Catalogue to retrieve uncharacterised GWAS loci SNPs in our meta-analysis and to get positional mapping information based on MAGMA^47^. We used a threshold of *P* < 1 x 10^-6^ to nominate tag SNPs. Additional SNPs that were in high LD with tag SNPs were inferred using European samples 1Kg Phase3 reference panel (with *r*2 > 0.6 and independent from each other with *r*2 < 0.6).

### Candidate gene analysis

In order to validate variants that have been reported in previous studies to be associated with time-to-LiD or LiD risk, we accessed the LiDPD website (Date accessed: 12/01/2023; http://LiDpd.eurac.edu/) and downloaded a list of curated variants from the literature. We explored these in our time-to-LiD GWAS meta-analysis ^48^.

### LiD prediction modelling

We used PRSice software (version 2; RRID:SCR_017057) to compute a polygenic risk score (PRS) using the summary statistics of our time-to-LiD meta-analysis as base data and the Tracking Parkinson’s cohort as target data. We chose the Tracking Parkinson’s cohort as it is the single largest cohort, which reduces the standard error (SE) of the PRS estimates, leading to more confident estimates. We then replicated the association of the nominated SNPs composing the PRS in the second largest cohort we had access to, OPDC, resembling a discovery / replication study design, although in this case the OPDC data had contributed to the LID PRS.

We set a threshold of *P* < 1 × 10^−6^ to nominate GWAS variants that make up the PRS. We selected independent SNPs by clumping within ±250 Kb from the index SNPs (the most significant SNP on a Kb window). We used the SNP betas as the estimated to compute the PRS from. Sex, standardised AAO, and the first 5 PCs were added as covariates to the PRS estimation process. To compute the LD estimates, we used the imputed cohorts from which we calculated the PRS, as they were large enough to provide accurate LD estimates (N > 500). To validate the PRS as an instrument to distinguish between PD patients with and without LiD, we derived time-dependent ROC curves, under the assumption that different PRS loads might cause changes to time-to-LiD onset. We used the Inverse Probability of Censoring Weighting (IPCW) estimation of Cumulative/Dynamic time-dependent ROC curve from the ‘timeROC’ R package (version 0.4; https://cran.r-project.org/web/packages/timeROC/index.html). To compute the weights, we used the Kaplan-Meier estimator of the censoring distribution.

Next, we used a stepwise logistic regression model with an in-house script using the ‘stats’ R base package (version 4.2.2; https://search.r-project.org/R/refmans/stats/html/00Index.html) to find whether any baseline clinical variable was significantly associated with LiD status. We used data from the Tracking Parkinson’s cohort, as it is deeply phenotypically characterised (number of baseline covariates = 702). After removing variables with high missingness rate (missing rate > 10%) or categorical variables with only one level, we defined a total of 502 baseline features (including the PRS) (eData 1). Then, we created a base logistic regression model (adjusted for sex the first 5 PCs and standardised AAO). At each step of the stepwise regression approach, we refitted the base model with each of the baseline predictors individually, and selected the model with the variable that decreased AIC the most. We ran the model until no variable further decreased the AIC, or until the AIC score was equal to 1. Once the model was fitted, we selected only those predictors that were significantly associated with the binary outcome, applying the conservative Bonferroni correction accounting for the number of predictors assessed. We set the significance threshold as 0.05 / 502 = 1 × 10^−4^.

## Results

### Cohort clinical features and prevalence

Across all cohorts (n= 2,784 PD patients), the rate of LiD was 14%, consistent across cohorts (Table 1), except in the PPMI cohort where it was 21%. This is consistent with other studies that reported that younger PD onset is more frequently associated with LiD^49–51^, given that PPMI is a *de novo* study that recruited younger patients on average. We did not exclude any patient from the PPMI cohort due to left-censoring.

**Table 1.**
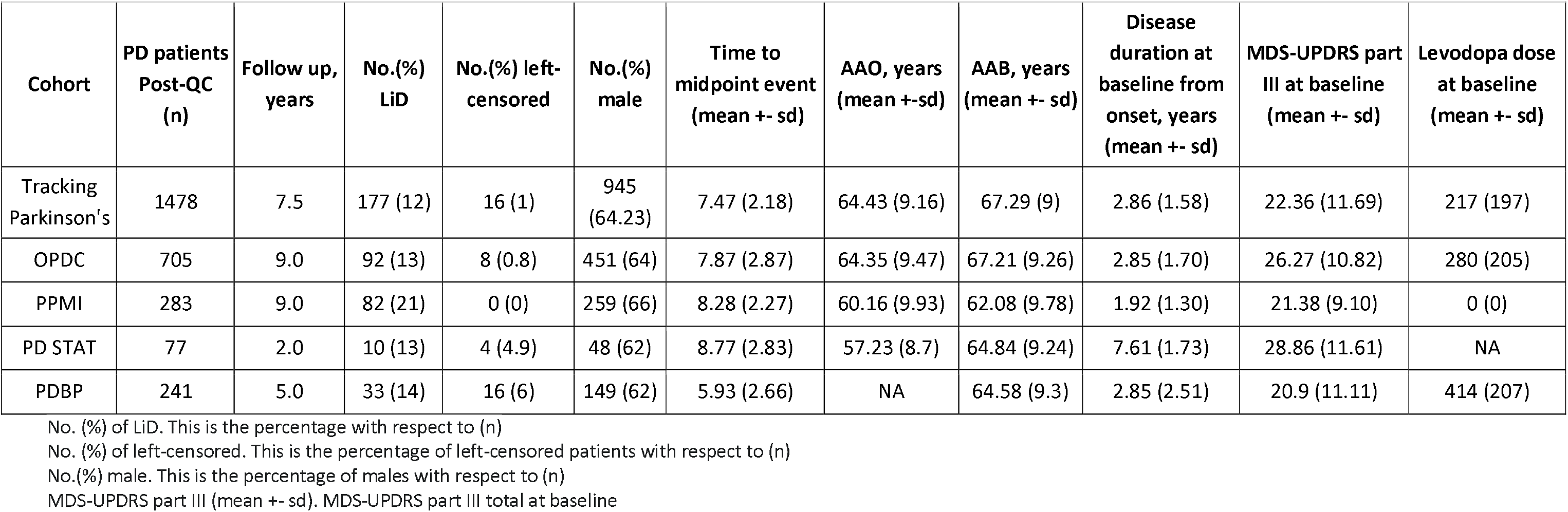
Cohort summary statistics.

We explored the effect of demographic and clinical factors previously reported to be associated with LiD. We merged baseline clinical data from all the cohorts. We found that patients with younger PD AAO (grouped as people with age at onset higher than 50 years and lower or equal than 50 years), had a higher probability of developing LiD than older patients along the time interval from disease onset to study end (HR = 1.8, SE = 0.14, *P* = 2 × 10^−5^) (data excluding PDBP as AAO was not available). Female PD patients showed a consistent decrease in the survival probability (increase in the probability of developing LiD) during a 12.5 years time interval (eFigure 2 a and b). Body mass index (BMI) was available in PPMI and Tracking Parkinson’s, and smoking status data was available in the Tracking Parkinson’s cohort only. We did not find a significant increase in the probability of developing dyskinesia either for PD patients with low baseline BMI nor for PD smokers at baseline (eFigure 2 c and d).

### Power analysis

We performed a power analysis to estimate the power to find a genetic association between time-to-LiD and genome-wide SNPs with the current sample size and LiD event rate, and how this varied with a range of genotype hazard ratios (GHRs) and AFs. We were well-powered (80% power) to detect genetic variants associated with the development of LiD with a HR equal or higher than 2 and a MAF as low as 0.01 (eFigure 3a). In addition, we performed a simulation to show as the sample size increases, the power to detect rarer associations improves. As we increased the sample size to 18000, we achieved 80% power for genetic variants with a MAF lower than 0.01, and with a HR lower than 2 (eFigure 3b).

### Time-to-LiD GWAS

We ran time-to-LiD GWAS independently for each cohort. We confirmed that there was no genomic inflation in any cohort-specific GWAS (eTable 3). We identified three loci significantly associated with time-to-LiD onset in the meta-analysis of the adjusted model on chromosome 1, chromosome 16 and chromosome 4 (Figure 1). The most significant SNPs at each loci were rs72673189, rs189093213, rs180924818. **rs72673189** (HR = 2.77, SE = 0.18, *P* = 1.53 × 10^−8^) in chromosome 1, is a variant in the third intron of the *LRP8* gene. **rs189093213** (HR = 3.06, SE = 0.19, *P* = 2.81 × 10^−9^) in chromosome 4 was found in the non-coding RNA *LINC02353 (PCDH7 1.2Mb downstream)*. **rs180924818** (HR = 3,13, SE = 0.20, *P* = 6.27 × 10^−9^) in chromosome 16 was found very close (0.15Mb upstream) to the 3’-UTR of the *XYLT1* protein coding gene in a non-coding region of the genome (Table 2). The direction of the effects was consistent and replicated across the meta-analysed cohorts in which the SNPs were present (Figure 2). To visually represent the survival probability of patients carrying the lead SNP on each locus we found on our meta-analysis, we extracted per cohort patients’ genotypes and showed the difference in the probability of LiD between carriers and non carriers through Kaplan-Meier curves (Figure 3).

**Figure 1.**
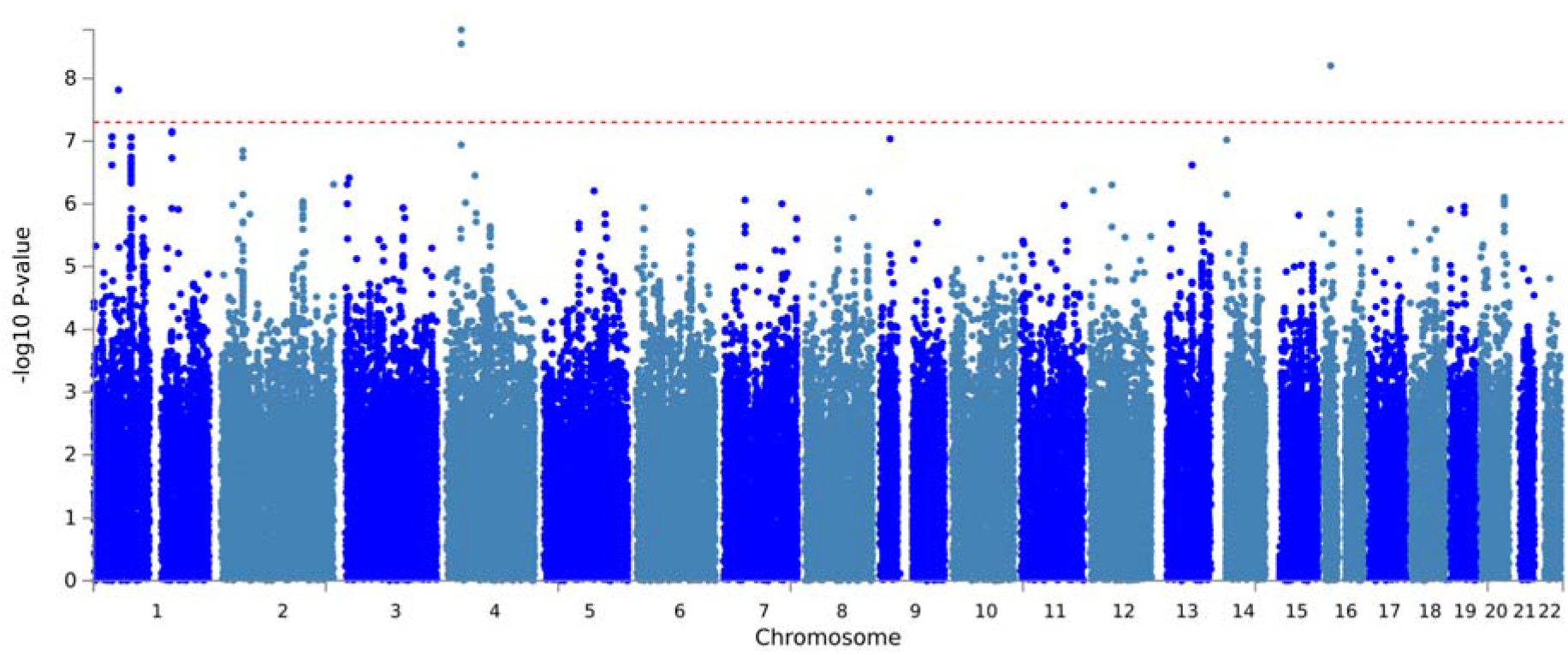
LiD GWSS meta-analysis Manhattan plot. The GWSS was conducted using a Cox proportional hazards model in each cohort separately, and results were meta-analysed. Red dots indicate the variant with the lowest *P*-value at each genome-wide significant genetic *locus*. Genome-wide significance was set at 5×10^-8^ and is indicated by the red dashed line.

**Figure 2.**
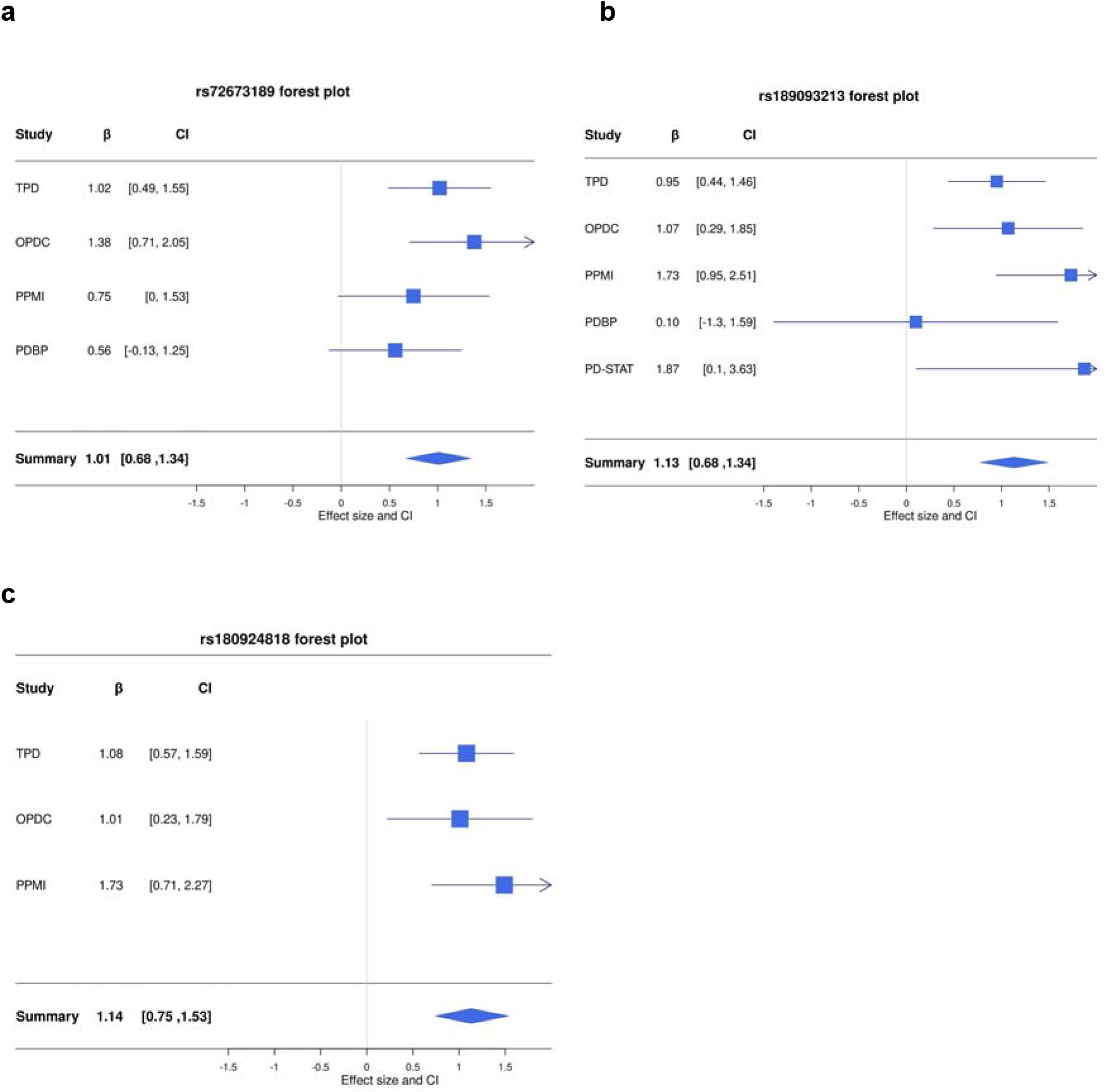
Forest plots of top hits of GWSS meta-analysis. **a**, *LRP8* rs72673189 variant (I² = 0; Cochran’s Q test: ꭓ^2^ = 0.24, df =, *P* = 1.53e-08). **b**, *LINC02353* rs189093213 variant (I² = 21.4; ꭓ^2^ = 5.09, df =, *P* = 1.67e-09). **c**, *LINC02353* rs180924818 variant (I² = 0; ꭓ^2^ = 0.77, df = 3, *P* = 6.27e-09). *HR* hazard ratio, *CI* confidence interval, *P p-*value, *r2* imputation info score.

**Figure 3.**
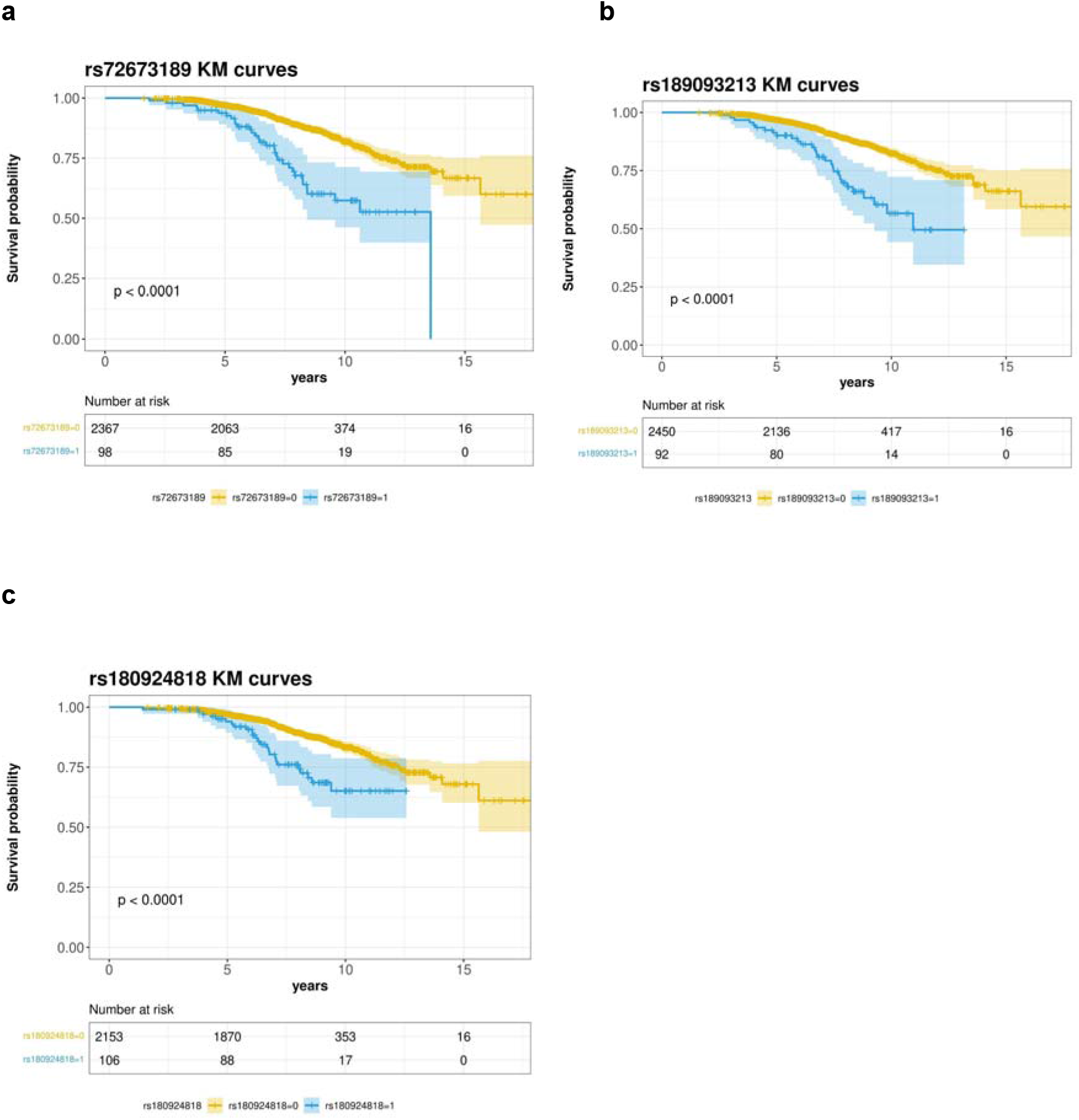
Survival curves of candidate SNPs. **a**, Kaplan-Meier curve for Survival probability (LiD free probability) based on rs72673189 carrier status in PD patients. **b**, Kaplan-Meier curve for Survival probability (LiD free probability) based on rs189093213 carrier status in PD patients. **c**, Kaplan-Meier curve for Survival probability (LiD free probability) based on rs180924818 carrier status in PD patients. The blue curve represents genetic variant carriers, whereas the yellow curve represents non-carriers. p = *p-*value. Number at risk represents the number of PD patients remaining on the study at the different time points (0, 5, 10, 15 years). The colour expansion on each curve represents the confidence interval (CI).

**Table 2.**
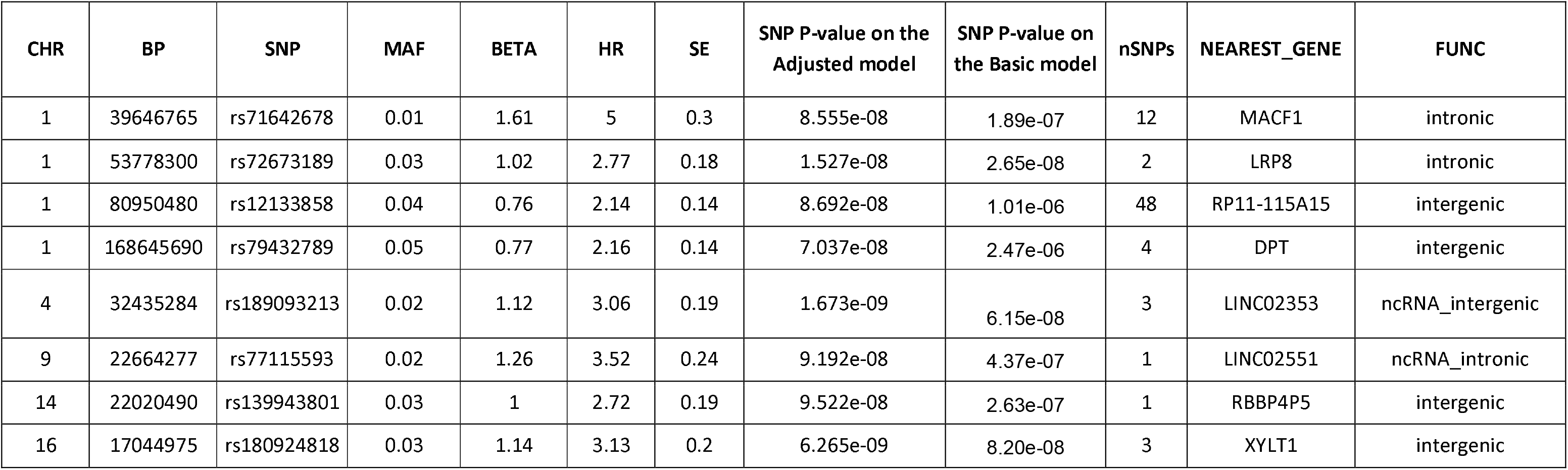
Independent significant SNPs with a P-value lower than 1e-7.

### Sensitivity analysis

The three variants found to significantly increase LiD susceptibility in the adjusted model approach remained associated in the basic model including only known confounders (eTable 4). We found the correlation of the SNP metrics between the basic and the adjusted model to be high (eFigure 4). This indicated that adding additional predictors based on baseline variation increased the power to detect SNP-outcome associations, presumably by explaining other sources of variance in the model, and that there was no source of confounding given by disease duration and severity measures (suggested by the high correlation in the SNP metrics).

Using data from Tracking Parkinson’s only, we investigated whether these associations could be confounded by levodopa dose or the disease stage at the LiD event time point. For each of the genome-wide significant SNPs, we repeated the CPH analysis adjusting for levodopa dose or disease stage as measured by MDS-UPDRS part III at the first visit when the LiD threshold was reached or at the last available visit for patients who did not develop LiD during the study length. We did not find a change either in the effect size or the test-statistics that could suggest an unaccounted source of confounding (eTable 4).

Finally, excluding PDBP from the meta-analysis did not have any significant change in the lead SNPs effect sizes and significance levels eTable 5).

### Functional annotation

We performed fine-mapping using ABF, SuSiE, FINEMAP, and Polyfun-SuSiE as described in Methods and found Consensus SNPs on each CPH GWAS nominated loci (eTable 6). We found the lead SNP on each locus to be Consensus SNPs, which are those selected by at least two different fine-mapping tools. We plotted each locus found to have at least one variant significantly associated with time to reach LiD against brain cell type-specific epigenomic data. We found that the lead (and fine-mapped SNP) at the *LRP8* locus belonged to a neuronal specific chromatin accessible region, which is a target region for DNA-associated proteins, as measured with the ATAC-seq and CHIP-seq (H3K27ac and H3K4me3) assays (Figure 4). We also found this SNP to be part of a neuronal specific enhancer-promoter interaction within *LRP8,* as defined by PLAC-seq (Figure 4). This implies that this specific *LRP8* intronic signal is an active neuronal enhancer of the *LRP8* expression, forming an anchored chromatin loop recruiting the transcription machinery to the *LRP8* transcription start site (TSS). In addition, we found suggestive evidence for the lead SNP lying in a transcription factor binding site (TFBS), as measured by the ENCODE project (eFigure 5). Similarly, we found that some of the fine-mapped SNPs (including the lead SNP) in the *XYLT1* locus were forming chromatin loops towards the *XYLT1* promoter, as measured by the PLAC-seq assay, suggesting that regulation of this gene associated with susceptibility to LiD (eFigure 6). We found this region to also overlap with TFBS marks (eFigure 7). We did not find any functional regulatory mark at the *LINC02353* locus.

**Figure 4.**
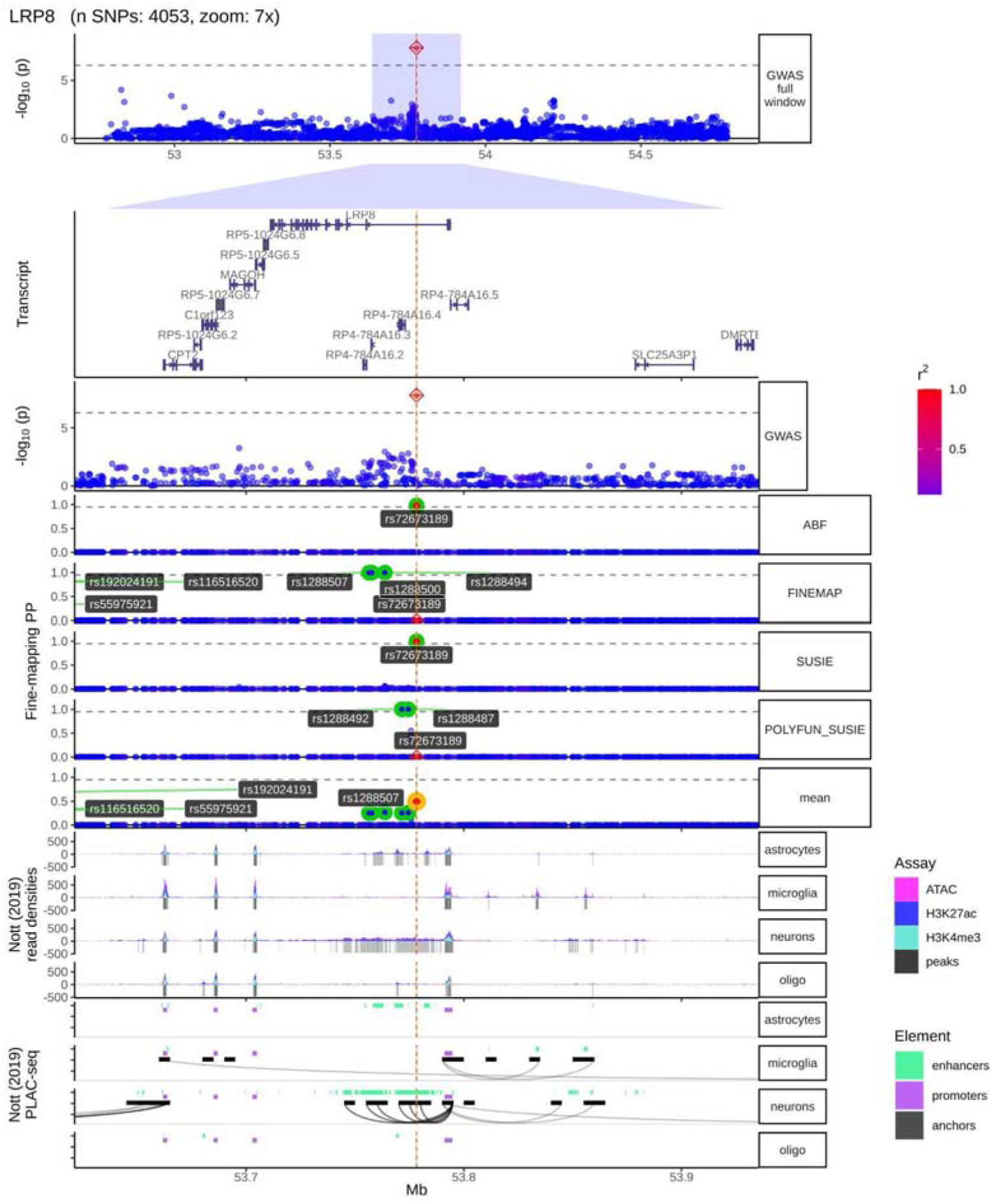
LRP8 locus fine-mapping and brain cell type specific regulatory marks. From top to bottom, locus plot, transcript plot, the fine-mapping nominated variants across fine-mapping tools, brain cell type specific regulatory element marks. In the locus plot, the SNPs are coloured in red as LD (given by R2) increases, and blue as the LD decreases. In the fine-mapping track, we highlight the SNPs with the highest posterior probabilities for each fine-mapping tool (ABF, FINEMAP, SUSIE, POLYFUN_SUSIE). In addition, we highlight in yellow the Consensus SNP with the highest mean Posterior Probability (mean). In the cell type specific regulatory element marks, the first 4 rows are the density marks (y-axis) from ATAC-seq assay (in pink), and CHIP-seq assays (H3K27ac in blue, and H3K4me3 in cyan), in astrocytes, microglia, neurons, and oligodendrocytes. The next four rows are the distal anchored chromatin loops (black curves). We see how, only in neurons, there is a chromatin loop forming from the *LRP8* GWS and the fine-mapped consensus variant towards the LRP8 promoter (purple).

Next, we performed colocalization analysis in all genes within 1Mb from lead SNPs with *P* < 1 × 10^−7^. We found suggestive support for colocalization between the LiD GWAS meta-analysis signals and ci-eQTL data from Metabrain Cortex (PP H4 > 0.7 on the unadjusted colocalization analysis; PP H4 > 0.5 on the colocalization analysis after adjusting the priors based on the number of overlapping SNPs in the locus of interest) for the *DNAJB4* gene on chromosome 1 (eTable 7). We did not find evidence of colocalization in the *XYLT1, LRP8 n*or the non-coding RNA loci.

A few loci approaching genome-wide significance (GWS) in chromosome 1, were in proximity with *DNAJB4*. Therefore, we decided to investigate if the single causal variant assumption holds in the *DNAJB4* locus, necessary to validate the colocalization signal in *DNAJB4*. We ran GCTA-COJO under stepwise and conditional model selection procedures. We filtered all SNPs within the *DNAJB4* locus that were used to perform the colocalization analysis and that matched the AMP-PD reference panel (4590 out of 4840 SNPs included in the colocalization analysis). After performing the stepwise selection procedure assuming complete linkage equilibrium between SNPs that are more than 10Mb from each other, and setting a collinearity cutoff of 0.9, only the lead SNP in the locus retained nominal significance (rs278853, MAF = 0.26, beta = 0.40, se = 0.08, *P* = 4.07 × 10^−6^). Similarly, running an association analysis on each of the 4590 SNPs conditioning on the lead variant (rs278853) did not show any of these SNPs to be nominally significantly associated, confirming the single causal variant assumption and that the results obtained with coloc on the *DNAJB4* locus were unbiased. Lastly, to understand whether the *DNAJB4* signal was independent of the GWS *LRP8* locus signal, we ran an analysis conditioning on the genome-wide significant *LRP8* SNP (rs72673189). We found that rs278853 remained nominally associated (*P* = 4.40 × 10^−6^), indicating these two signals were independently associated with the risk of developing LiD.

### Candidate variant analysis

We determined whether previously reported variants in the LiD literature (from LiDPD) had an impact on the time to LiD (eTable 8). We found *ANNK1* and *BDNF* variants to be nominally significantly associated (*P* < 0.05) with the time to dyskinesia. Nonetheless, ANNK1 or BDNF variants did not reach the significance threshold after applying Bonferroni correction according to the number of SNPs tested (*P* < 2 × 10^−3^).

*LRP8* has previously been reported to be associated with *APOE* and the microtubule associated protein tau (*MAPT*)^52^. A previous retrospective study including 855 caucasian PD patients found a suggestive association between the H1b *MAPT* haplotype and a higher likelihood of dyskinesia at an initial visit^53^. In the case of *XYLT1*, a previous study has found a regulatory effect of a *XYLT1* variant on the mRNA levels of *GBA* in the substantia nigra and cortex ^54^. We investigated whether MAPT variants (rs1800547; rs242562; rs3785883; rs2435207) were associated with the time to LiD. In addition, we explored whether *APOE* and *GBA* variants increased the risk to develop LiD ^55^. We did not find an association between time to LiD and *APOE* variants rs429358 and rs7412, or *GBA* rs2230288 variant (E326K), or MAPT rs1800547, rs242562, rs3785883, rs2435207.

In addition, we explored genetic associations from *PINK1*, *DJ-1,* and *PRKN* intergenic variants. Whereas we did not find any genetic variant associated with time to LiD on the PINK1 locus, we found 26 *DJ-1* intergenic variants on the with a P-value < 0.05 (rs1641433611 lead SNP; HR = 1.84, SE = 0.2, *P* = 4 × 10^−4^). Similarly, we found 162 intergenic variants with a Pvalue < 0.05 in the PRKN locus (rs113276175 lead SNP; HR = 1.84, SE = 0.2, *P* = 4 × 10^−4^) (eTable 9).

### PRS is capable of distinguishing patients that develop LiD

We nominated a total of 67 independent SNPs to compute the PRS in the Tracking Parkinson’s cohort (eTable 10). We then validated the proposed SNP set on the OPDC cohort by measuring the ability to distinguish LiD PD patients. We found that genetic data as summarised by PRS, without any other clinical or demographic data, could accurately distinguish PD patients that developed LiD at 10 years from disease onset in two separate cohorts: Tracking Parkinson’s (AUC 83.9) and OPDC (AUC 87.8) (eFigure 8).

### Stepwise regression approach to determine baseline predictors of LiD development

We used Tracking Parkinson’s data at baseline in a stepwise regression approach using a logistic model. We then filtered out from the final model predictors that were not significantly associated after applying Bonferroni correction (*P* < 0.05 / 502 = 1 × 10^−4^).

In addition to the PRS, which was significantly associated with a increase of the odds of LiD (OR = 962.94, SE = 0.57, *P* = 1.07 × 10^−30^), we found that anxiety at baseline (as measured by the Leeds Anxiety and Depression Scale^56^) was significantly associated with a increase of the odds of LiD (OR = 1.14, SE = 0.03, *P* = 7.4 × 10^−5^). We also explored clinical features previously reported as being associated with an increased or decreased LiD risk. Sex, AAO, and 5PCs were added in the base model of the stepwise regression approach. Consistent with previous studies as well as with our CPH model highlighted above, younger AAO increased the LiD odds (OR = 2.41, SE = 0.04, *P* = 4 × 10^−3^). However, sex was not found to be significantly associated in our final model including PRS and Leeds anxiety status.

Neither smoking status nor BMI were selected on the stepwise regression approach, consistent with what we found when we individually explored known LiD risk factors (eFigure 2). Interestingly, we also found that PD family history was selected on the stepwise regression analysis, and was nominally significantly associated with an increase in the odds of LiD (OR = 1.62, SE = 0.14, *P* = 6.9 × 10^−4^).

Finally, we attempted to replicate the association between dyskinesia state and anxiety using State-Trait Anxiety Inventory^57^ available in PPMI. We did not find the Trait Anxiety Score to be significantly associated with LiD patients in PPMI (OR = -0.03, SE = 0.04, *P* = 0.44).

### Patients with LiD have an average higher cognitive scoring

We were interested in assessing the cognitive status of LiD patients because of the association between the *LRP8* nominated locus and *APOE*. We explored whether the cognitive state differed between patients developing LiD and patients who did not develop LiD during the study length using the Wilcoxon rank sum non-parametric test with continuity correction, as we observed the data was not normally distributed. In addition, we also looked into differences in the MDS-UPDRS part III scores between the two groups, using the unpaired two samples t-test to compare the mean of two independent groups. We compared the LiD group (N = 172) against the non-LiD PD group (N = 1318) using data from Tracking Parkinson’s alone as it is the largest deeply phenotype cohort we had available. We did not find differences in the average MDS-UPDRS part III total score, either at baseline nor at the visit when patients first developed LiD (or the last available visit in cases who did not develop LiD) (eTable 11). On the other hand, we found that PD patients who did not develop LiD had a significantly lower MoCA score on average at baseline as well as at the visit the outcome was met (eTable 11).

## Discussion

We have performed an untargeted genome-wide study to uncover genetic variants associated with the time-to-LiD in PD, using a CPH model under a genetic additive effect and analysed the effect of genetic and baseline clinical variation on the development of LiD. We found genome-wide significant associations with the time-to-develop LiD at the *LRP8*, *LINC02353* and *XYLT1* loci. These associations were replicated across all the cohorts included in the meta-analysis. We also performed a candidate gene analysis, exploring genetic variants reported to be associated with LiD risk in our large GWAS meta-analysis. We found that genetic variability in *BDNF* and *ANKK2*, were nominally associated with LiD.

Post-hoc functional annotation analysis revealed a chromatin loop between an enhancer within *LRP8* third intron (where the lead variant was found) and the *LRP8* promoter, thus providing functional support for *LRP8* as the causal gene at this locus. In addition, a colocalization analysis, looking at all genes within ±1Mb from all GWAS variants with P-value < 1 × 10^−7^ revealed a second association in chromosome 1 with the *DNAJB4* gene. Conditional analysis further confirmed that both regions were in linkage equilibrium, hence both *LRP8* and *DNAJB4* were independently associated with the time-to-LiD. We also found a similar event of distal regulation in the *XYLT1* locus, although the chromatin loop did not perfectly match with the GWAS signals, making the functional annotation analysis inconclusive. Moreover, we found that the two GWAS nominated signals overlapped with Transcription Factor Binding Sites marks from the ENCODE project, adding further support for the transcription machinery being recruited in the GWAS loci and regulating both genes expression after forming the enhancer-promoter distal chromatin loops. Nevertheless, whereas we found a chromatin loop suggesting regulation of *XYLT1* and LRP8 gene expression, we did not find statistical support for gene regulation based on the colocalization Bayesian framework.

The three nominated protein coding genes have been previously reported to be functionally associated with putative PD genes, which may provide an insight into the development of LiD. *LRP8* encodes the low-density lipoprotein receptor-related protein 8, and it has been found to be associated with *APOE*. In addition, the *LRP8* protein stabilises microtubule associated protein tau *(MAPT*) and it has been shown that knocking out *LRP8* in mice increases tau phosphorylation^52^. DNAJB4 gene encodes a molecular chaperone tumour suppressor, and member of the heat shock protein-40 family. Mutations in the DNAJ family protein have been reported to cause or increase the risk of several neurological disorders, including Parkinson’s disease ^58^. In the case of *XYLT1*, it encodes a xylosyltransferase enzyme which takes part in the biosynthesis of glycosaminoglycan chains. A previous study has found a regulatory effect of a *XYLT1* variant on the mRNA levels of *GBA* in the substantia nigra and cortex ^54^. We did not find support for colocalization with eQTLs nor evidence suggestive of epigenetic regulation of genes in the *LINC02353* locus. *PCDH7*, the nearest gene coding protein gene, encodes a protein with an extracellular domain containing 7 cadherin repeats. This gene has been described as a potential PD biomarker ^59^.

At an individual patient level, treatment strategies including levodopa and non-levodopa therapies, and the use of deep brain stimulation (DBS) are determined by the emergence of motor complications including LID. The ability to develop a predictive algorithm to enhance clinical care would improve the outlook for PD treatment. Here, we have shown that both clinical and genetic variables have the potential to have a high predictive value for the development of LID. This will need to be validated in further cohorts and we hypothesise that the integration of further omics data (e.g. RNA and proteomics), using machine learning may lead to discovering an accurate predictive model determining PD patients at risk of developing dyskinesia when treated with the therapies available to date.

To our knowledge, this is the largest study to date with detailed clinical, drug exposure and genetic data. We have carefully tested for confounding by PD age at onset, gender, population structure and shown that our results are free of confounding effects as well as demonstrating they are consistent across cohorts. Because the dose of levodopa may be a major confounder in our study, we tested the effects of adjusting for levodopa dose on a sensitivity analysis, and found that the lead SNPs on *LRP8*, *LINC02353* and *XYLT1* loci remained significantly associated with the outcome, concluding that levodopa treatment was not a confounder in our study design. Likewise, adjusting for the MDS-UPDRS part III total score at the time of LiD development did not change the significance levels of the lead SNPs, suggesting that our findings were not confounded by the motor severity of the disorder. Although this is a large study there are limitations based on sample size. According to our sample calculation, we would be 80% powered to detect associations with the LiD phenotype from variants with a MAF of 0.01 when we reached a sample size of 18000 patients. Expanding this analysis on growing PD genetic datasets with deeply phenotypic data available from initiatives such as the Global Parkinson’s Genetic Program (GP2) will give us new insight into the genetics of PD LiD patients as well as serve as a valuable resource for validation of findings ^60^.

Overall, we have found new evidence of common genetic variability associated with the time-to-LiD. We have been able to map genes nearby risk loci, as well as give fine mapping support of which might be the causal variants of the LiD trait. Likewise, we hope to help design personalised medicine strategies that prevent PD patients developing dyskinesia according to their genetic burden which could be tested with the proposed PRS in this study. Similarly, we hope to help understand the molecular pathways that, when altered, lead to LiD. Targeting nominated genes might allow the development of LiD treatment strategies. Further investigation regarding the overlap between anxiety GWAS and our GWAS might help understanding common causal pathways between the two conditions. Understanding shared mechanisms will help us prevent medication adverse events affecting non-targeted pathways and to fine-tune current treatments.

## Supporting information

SUPPLEMENT_LID

## Data Availability

All data produced are available online at https://doi.org/10.5281/zenodo.7795604

https://doi.org/10.5281/zenodo.7795604

## Code availability

All the code has been made publicly available on GitHub (https://github.com/AMCalejandro/LID-CPH.git) DOI: https://doi.org/10.5281/zenodo.7802142) Analyses were performed using open-source tools as described in the Methods section.

## Data availability

GWAS summary statistics are publicly available in the Zenodo ASAP data repository (https://doi.org/10.5281/zenodo.7795604). Supplementary Figures and Tables are available in the Zenodo ASAP data repository (https://zenodo.org/record/7802755#.ZC2RAnbMK38). TPD data is available upon access request from https://www.trackingparkinsons.org.uk/about-1/data/. The PDBP and PPMI data was accessed from Accelerating Medicines Partnership: Parkinson’s Disease (AMP-PD) and data is available upon registration at https://www.amp-pd.org/. OPDC data is available upon request from the Dementias Platform UK (https://portal.dementiasplatform.uk/Apply). PD-STAT is available upon request to the principal investigator (C Carroll, Plymouth University, https://penctu.psmd.plymouth.ac.uk/pdstat/#:~:text=PD%20STAT%20%2D%20Simvastatin%20as%20a,brain%20from%20injury%20or%20loss.). HapMap phase 3 data (HapMap3) is available for download at ftp://ftp.ncbi.nlm.nih.gov/hapmap/. Cis-QTL eQTLGen data was downloaded from (https://www.eqtlgen.org/cis-eqtls.html). MetaBrain cis-eQTL data can be accessed upon access request form (https://www.metabrain.nl/cis-eqtls.html). eQTL data from eQTL catalogue can be ftp-accessed (https://www.ebi.ac.uk/eqtl/Data_access/). ENCODE TFBS marks and Nott brain cell type-specific enhancer-promoter interactome data were accessed through echolocatoR. (https://github.com/RajLabMSSM/echolocatoR).

## Acknowledgements

This research was funded in whole or in part by Aligning Science Across Parkinson’s [Grant number: ASAP-000478] through the Michael J. Fox Foundation for Parkinson’s Research (MJFF). For the purpose of open access, the author has applied a CC BY public copyright licence to all Author Accepted Manuscripts arising from this submission.

This research was supported by the National Institute for Health Research University College London Hospitals Biomedical Research Centre. The UCL Movement Disorders Centre is supported by the Edmond J. Safra Philanthropic Foundation. This work was supported in part by the Intramural Research Program of the National Institute on Aging (NIA).

Data used in the preparation of this article were obtained from the AMP-PD Knowledge Platform (https://www.amp-pd.org). AMP-PD is a public-private partnership managed by the FNIH and funded by Celgene, GSK, Michael J. Fox Foundation for Parkinson’s Research, the National Institute of Neurological Disorders and Stroke, Pfizer, and Verily. Clinical data and biosamples used in preparation of this article were obtained from the Parkinson’s Progression Markers Initiative (PPMI), and the Parkinson’s Disease Biomarkers Program (PDBP).

PPMI – a public-private partnership – is funded by the Michael J. Fox Foundation for Parkinson’s Research and funding partners, including [list the full names of all of the PPMI funding partners found at https://www.ppmi-info.org/about-ppmi/who-we-are/study-sponsors]. The PPMI Investigators have not participated in reviewing the data analysis or content of the manuscript. For up-to-date information on the study, visit www.ppmi-info.org.

The Parkinson’s Disease Biomarker Program (PDBP) consortium is supported by the National Institute of Neurological Disorders and Stroke (NINDS) at the National Institutes of Health. A full list of PDBP investigators can be found at https://pdbp.ninds.nih.gov/policy. The PDBP Investigators have not participated in reviewing the data analysis or content of the manuscript.

Both TPD and OPDC cohorts are primarily funded and supported by Parkinson’s UK (https://www.parkinsons.org.uk/) and supported by the National Institute for Health and Care Research (NIHR) Clinical Research Network (CRN). The TPD study is also supported by NHS Greater Glasgow and Clyde. The OPDC cohort is also supported by the NIHR Oxford Biomedical Research Centre, based at the Oxford University Hospitals NHS Trust, and the University of Oxford. PD-STAT is funded and supported by grants from the Cure Parkinson’s Trust (https://cureparkinsons.org.uk/) and JP Moulton Charitable Foundation (https://www.perscitusllp.com/moulton-charity-trust/), co-ordinated by the Peninsula Clinical Trials Unit, University of Plymouth and sponsored by University Hospitals Plymouth NHS Trust.

## Author Contributions

H.R.M. and A.M.C designed the study. H.R.M. supervised the study. D.G.G., M.T.M.H., Y.B.-S., M.A.L,, J.H. and H.R.M. conceived and led the TPD and OPDC clinical cohorts, as well as performed data management and curation. A.M.C performed all the quality control on each cohort and performed all analyses in the present manuscript. C.C. led the PD STAT study, as well as performed data management and curation. M.M.X.T. helped with the design of the quality control strategy. L.W provided access to an harmonised version of the AMP-PD genetic data. M.A.L helped with the statistical study design. M.S helped to design the statistical and quality control methodology. H.I provided the base code to run a stepwise regression approach for clinical features selection. T.F reviewed the manuscript methodology and results. A.M.C wrote the initial manuscript. All authors critically reviewed the manuscript

