## SUPPLEMENT_LID for "Genetic meta-analysis of levodopa induced dyskinesia in Parkinson’s disease"

#### **Supplementary Online Content**

**1. eMethods**

1.1 Genotype data quality control and imputation

1.2 Time to LiD midpoint

1.3 Colocalization analysis

1.4 Sensitivity analyses

**2. eTables**

**eTable 1.** Study sample sizes and genotyping array.

**eTable 2.** List of covariates added on both the basic and adjusted model

across cohorts

**eTable 3.** Genomic inflation values in each cohort and meta-analysis

summary statistics

**eTable 4.** Sensitivity analyses lead SNP P-values in the basic CPH model for the TPD cohort

**eTable 5.** Lead SNP P-values in the CPH model including and excluding PDBP cohort in the basic and adjusted models

**eTable 6.** List of fine-mapped consensus SNPs on each locus.

**eTable 7.** Colocalization hypotheses posterior probabilities

**eTable 8.** Candidate variants analysis.

**eTable9.** List of DJ-1, and PARKIN intergenic variants with a significance value lower than 0.05 on the raw GWAS meta-analysis

**eTable 10.** Table of SNPs used to derive the GRS in the TPD cohort

**eTable 11.** MoCa and UPDRS score comparison between PD-LiD and PD groups

**3. eFigures**

**eFigure 1.** Quality control flowchart.

**eFigure 2.** LiD risk factors Kaplan Meyer curves.

**eFigure 3**. Power calculation and simulation.

**eFigure 4**. SNP metrics correlation between the CPH basic and best models.

**eFigure 5.** LRP8 locus fine-mapping and top 5 TFBS marks.

**eFigure 6.** XYLT1 locus fine-mapping and brain cell type specific regulatory marks.

**eFigure 7.** XYLT1 locus fine-mapping and top 5 TFBS marks.

**eFigure 8.** PRS Receiver operating characteristic (ROC) curves.

**4. eData**

**eData 1.** List of all covariates screened in the stepwise regression model

**5. eReferences**

**1. eMethods. Genotype data quality control and imputation, Time to LiD midpoint, Colocalization analysis, Sensitivity analyses**

**1.1. Genotype data quality control and imputation**

**Sample QC**: Using PLINK v1.9 we used X chromosome genotype data to check for sex discordance between the genotypic and phenotypic sex. We excluded individuals who were missing more than 5% of genotypes. Samples with excess or reduced heterozygosity in autosomes (defined as ± 4 standard deviations (SD) away from the mean heterozygosity rate within each cohort) were also excluded, as it can indicate contamination or increased homozygosity, respectively. We removed related individuals. Using GCTA software (version 1.93.0 beta for Linux; <https://yanglab.westlake.edu.cn/software/gcta/#Overview>)^1^, we created a relationship matrix from pruned genotypes, and we filtered out individuals which had a similarity score higher than 0.125, equivalent to 1st degree relatives. Population stratification is a major confounder in genetic association studies due to differences in allele frequencies between ethnicity groups. We therefore excluded individuals of non-European ancestry by performing principal component analysis (PCA) using the HapMap reference panel (Release number 3; ftp://ftp.ncbi.nlm.nih.gov/hapmap/)^2^. Individuals who were 6 SD away from the Northern and Western European ancestry (CEU) sample mean for any of the first 10 principal components (PCs) were considered ancestry outliers and removed.

**Variant QC**: PLINK v1.9, at a genotype level, we removed variants that had a missing rate higher than 0.05, variants with a minor allele frequency (MAF) of less than 0.01, and variants in which missing calls were not randomly distributed, by testing whether missingness status could be predicted from genotype calls at the two adjacent variants. We excluded variants that deviated from Hardy-Weinberg equilibrium (HWE), as extreme HWE deviations can be indicative of genotyping errors (*P* < 1 × 10^−10^)^3^.

**Imputation and post-imputation QC**: We ran the Will Rayner tool (Version 4.2.10; <https://www.well.ox.ac.uk/~wrayner/tools/>) for further quality checks against the Haplotype Reference Consortium (HRC) (GRCh37/hg19) panel (version r1.1 2016; <http://www.haplotype-reference-consortium.org/site>). Likewise, we updated strand, position, and reference / alternate allele assignment, as well as removed A/T and G/C SNPs if MAF> 0.4, SNPs with allele frequency difference > 0.2 compared to the reference panel, and SNPs not present in the HRC Panel ^4^. We then imputed each QCed cohort in the Michigan Imputation Server (RRID:SCR_017579; <https://imputationserver.sph.umich.edu>)^5^ using Minimac4 ( version 1.0.0; RRID:SCR_009292 [https://genome.sph.umich.edu/wiki/Minimac4 version 1.0.0](https://genome.sph.umich.edu/wiki/Minimac4)) and Eagle2 (v2.4; RRID:SCR_017262 ) with 20-Mb chunk sizes used to estimate haplotype phasing. We used the HRC panel as the reference panel of individuals of predominantly European ancestry for imputation. Once the data was imputed, we filtered out variants with a Rsq score < 0.7, to preserve only the variants imputed with high confidence. Finally, we removed variants with missingness rate > 5% and MAF < 1%.

**1.2.Time to LiD midpoint**

Because the precise time patients first developed dyskinesia happens between the visit in which dyskinesia were first recorded and the previous visit, we set the time to develop dyskinesia as the midpoint between the last visit with no LiD and the first visit with LiD. We set age at motor onset as the start point to measure time to develop LiD. Patients who did not develop LiD at the end of the study or at the time of withdrawal were right-censored. In patients who withdrew from the study and where withdrawal time was not available, we set the censoring time as the midpoint between the last visit patients attended clinic and the next scheduled visit.

**1.3. Colocalization analysis**

We evaluated all genes within ± 1Mb from the lead variants with a *P* <  1 × 10^−7^ at each GWAS locus ^6^. Coloc makes use of Bayesian inference to compute the posterior probability (PP) of five different hypothesis: No association with either trait (H0); Association with the LiD trait but not the eQTL trait (H1); Association with the eQTL trait but not the LiD trait (H2); Association with both traits, but the causal variant is distinct (H3); and the that there is a shared causal variant associated with both traits (H4). Each PP hypothesis lies between 0 and 1 and a high PPH4 (PPH4 > 0.8) is considered as evidence of colocalization between two traits tested, meaning the GWAS variant causes changes in specific gene expression. We ran coloc using default p_1_ = 1 × 10^−4^, p_2_ = 1 × 10^−4^, and p_12_ = 1 × 10^−5^ priors (p_1_ and p_2_ are the prior probability that any random SNP in the region is associated with trait 1 and 2, respectively, while p_12_ is the prior probability that any random SNP in the region is associated with both traits). However, it is worth noting that the prior for H3 hypothesis (association with both phenotypic and expression traits, but distinct causal variants) is ≈ *n*(*n* − 1)*p*1.*p*2, which scales with the square of *n*, resulting in H3 becoming more likely than H4 as the number of overlapping SNPs in the region increases^7^. Therefore, we adjusted the priors to account for the high number of overlapping SNPs (p_1_ = 3 × 10^−5^, p_2_ =  3 × 10^−5^, and p_12_ = 5 x 10^−7^ )^8^.

**1.4. Sensitivity analyses**

We performed four sensitivity analyses to assess whether the best model described above led to an unbiased testing of the null hypothesis of no association between all genome-wide SNPs and time-to-LiD. The first sensitivity analysis was designed to compare the basic and adjusted models. We tested whether high deviations in the SNP estimates and P-values arose after accounting for disease severity and dopaminergic denervation at baseline by measuring the correlation between the basic and adjusted GWAS meta-analyses. Next, we performed two separate sensitivity analyses to test whether either levodopa dose or the PD motor severity (as measured by MDS-UPDRS part III) at the time point where LiD were first documented, were confounding our findings. We performed this sensitivity analysis in Tracking Parkinson's, the largest dataset. We performed a CPH GWAS on the Tracking Parkinson's cohort adjusting by: a) known confounders, b) known confounders + motor severity (as measured by MDS-UPDRS part III) c) known confounders + levodopa dose. We compared the SNP metrics from the three models for the lead SNPs on the loci that reached genome-wide significance on the time-to-LiD GWAS meta-analysis. Lastly, because the PDBP cohort did not have age at onset available and we used AAD in the CPH model, we reran the time-to-LiD GWAS meta-analysis excluding PDBP to confirm that this cohort was not inflating the SNP test-statistics.

**2. eTables.**

**eTable 1. Study sample sizes and genotyping array.**

| **Study Name** | **Samples source** | **Abbreviations** | **N** | **Genotyping array** |
| --- | --- | --- | --- | --- |
| Tracking Parkinson’s Disease | UK | TPD | 2000 | Illumina HumanCoreExome array |
| Oxford Parkinson's Disease Centre Discovery Cohort | UK | OPDC | 1082 | Illumina HumanCoreExome-12 v1.1 or  Illumina Infinium HumanCoreExome-24 v1.1 |
| Parkinson's Progression Markers Initiative | USA | PPMI | 415 | WGS |
| Advancing Parkinson’s Disease Biomarkers Discovery | USA | PDBP | 873 | WGS |
| Simvastatin as a neuroprotective treatment for Parkinson’s disease | UK | PD-STAT | 174 | Illumina Neurochip |

WGS = Whole Genome Sequencing

**eTable 2. List of covariates added on both the basic and adjusted model across cohorts**

| **Study Name** | **Covariates in basic model** | **Covariates in the adjusted model** |
| --- | --- | --- |
| Tracking Parkinson’s Disease | AAO, GENDER, 5 PCs | AAO, GENDER, 5 PCs, BASELINE DISEASE DURATION, BASELINE MDS-UPDRS-III total, BASELINE L-DOPA DOSE |
| Oxford Parkinson's Disease Centre Discovery Cohort | AAO, GENDER, 5 PCs | AAO, GENDER, 5 PCs, BASELINE MDS-UPDRS-III total, BASELINE LEDD |
| Parkinson's Progression Markers Initiative | AAO, GENDER, 5 PCs | AAO, GENDER, 5 PCs, BASELINE MDS-UPDRS-III total, BASELINE DISEASE DURATION |
| Advancing Parkinson’s Disease Biomarkers Discovery | AAD, GENDER, 5 PCs | AAD, GENDER, 5 PCs, BASELINE MDS-UPDRS III total, BASELINE DISEASE DURATION |
| Simvastatin as a neuroprotective treatment for Parkinson’s disease | AAO, GENDER, 5 PCs | AAO, GENDER, 5 PCs |

**eTable 3. Genomic inflation values in each cohort and meta-analysis summary statistics**

| **Cohort Name** | **Lambda** |
| --- | --- |
| TPD | 1.02 |
| OPDC | 0.99 |
| PPMI | 0.97 |
| PDBP | 0.87 |
| PD-STAT | 0.73 |
| META-ANALYSIS | 1.04 |

**eTable 4. Sensitivity analyses lead SNP P-values in the basic CPH model for the TPD cohort**

| **CHR** | **BP** | **SNP** | **A1** | **A2** | **Basic model P-value** | **Levodopa model P-value** | **MDS-UPDRS III model P-value** |
| --- | --- | --- | --- | --- | --- | --- | --- |
| 1 | 53778300 | rs72673189 | A | G | 1.96E-04 | 2.39E-04 | 2.50E-04 |
| 4 | 32435284 | rs189093213 | A | G | 6.32E-03 | 1.89E-03 | 3.49E-03 |
| 16 | 17044975 | rs180924818 | G | A | 1.69E-04 | 2.62E-04 | 1.21E-04 |

Basic model P-value = P-value on the basic model without confounding variables

Levodopa model P-value = Levodopa dose at the time point the outcome was met

MDS-UPDRS III model P-value = MDS-UPDRS III total score at the time point the outcome was met.

**eTable 5. Lead SNP P-values in the CPH model including and excluding PDBP cohort in the basic and adjusted models**

| **CHR:POS** | **SNP** | **A1** | **A2** | **MAF** | **BETA** | **SE** | **P-value** | **N** | **PDBP INCLUDED** | **MODEL** |
| --- | --- | --- | --- | --- | --- | --- | --- | --- | --- | --- |
| 4:32435284 | rs189093213 | A | G | 0.02 | 1.1245 | 0.19 | 1.673e-09 | 2687 | YES | ADJUSTED |
| 4:32435284 | rs189093213 | A | G | 0.02 | 1.0032 | 0.18 | 6.154e-08 | 2784 | YES | BASIC |
| 4:32435284 | rs189093213 | A | G | 0.02 | 1.1894 | 0.19 | 6.24e-10 | 2446 | NO | ADJUSTED |
| 4:32435284 | rs189093213 | A | G | 0.02 | 1.0594 | 0.19 | 2.989e-08 | 2543 | NO | BASIC |
| 16:17044975 | rs180924818 | A | G | 0.98 | -1.14 | 0.20 | 6.265e-09 | 2687 | YES | ADJUSTED |
| 16:17044975 | rs180924818 | A | G | 0.98 | -1.037 | 0.19 | 8.197e-08 | 2784 | YES | BASIC |
| 16:17044975 | rs180924818 | A | G | 0.98 | -1.14 | 0.20 | 6.265e-09 | 2446 | NO | ADJUSTED |
| 16:17044975 | rs180924818 | A | G | 0.98 | -1.037 | 0.19 | 8.197e-08 | 2543 | NO | BASIC |
| 1:53778300 | rs72673189 | A | G | 0.0 | 1.0163 | 0.18 | 1.527e-08 | 2610 | YES | ADJUSTED |
| 1:53778300 | rs72673189 | A | G | 0.02 | 1.0005 | 0.18 | 2.654e-08 | 2707 | YES | BASIC |
| 1:53778300 | rs72673189 | A | G | 0.02 | 1.0743 | 0.19 | 1.505e-08 | 2369 | NO | ADJUSTED |
| 1:53778300 | rs72673189 | A | G | 0.02 | 1.0411 | 0.19 | 4.214e-08 | 2466 | NO | BASIC |

**eTable 6. List of fine-mapped consensus SNPs on each locus.**

| **SNP** | **CHR** | **P-value** | **Effect** | **SE** | **A1** | **A2** | **MAF** | **leadSNP** | **ABF.PP** | **FINEMAP.PP** | **SUSIE.PP** | **POLYFUN_SUSIE.PP** | **Support** | **mean.PP** | **mean.CS** | **Locus** |
| --- | --- | --- | --- | --- | --- | --- | --- | --- | --- | --- | --- | --- | --- | --- | --- | --- |
| rs72673189 | 1 | 1.5e-08 | 1.01 | 0.18 | A | G | 0.02 | TRUE | 1 | 0 | 1 | 0 | 2 | 0.50 | 0 | LRP8 |
| rs180924818 | 16 | 6.2e-09 | -1.14 | 0.20 | A | G | 0.02 | TRUE | 1 | 1 | 1 | 1 | 4 | 1 | 1 | LXYLT1 |
| rs137895239 | 16 | 3.1e-05 | 0.88 | 0.21 | A | G | 0.02 | FALSE | 0 | 1 | 1 | 1 | 3 | 0.75 | 0 | LXYLT1 |
| rs142441980 | 16 | 1.4e-06 | 0.88 | 0.18 | A | G | 0.03 | FALSE | 0.01 | 1 | 1 | 1 | 3 | 0.75 | 0 | LXYLT1 |
| rs17207399 | 16 | 2e-04 | -0.47 | 0.13 | C | G | 0.07 | FALSE | 1 | 1 | 1 | 1 | 3 | 0.75 | 0 | LXYLT1 |
| rs189093213 | 4 | 1.7e-09 | 1.12 | 0.19 | A | G | 0.01 | TRUE | 0.61 | 0.96 | 1 | 1 | 3 | 0.90 | 0 | LINC02353 |
| rs10023843 | 4 | 0.1 | 0.55 | 0.36 | T | G | 0.02 | FALSE | 0 | 0 | 1 | 1 | 2 | 0.50 | 0 | LINC02353 |
| rs139511855 | 4 | 4.5e-05 | -1.09 | 0.27 | A | G | 0.01 | FALSE | 0 | 0 | 1 | 1 | 2 | 0.50 | 0 | LINC02353 |
| rs147573196 | 4 | 2.5e-06 | 1.20 | 0.25 | A | T | 0.01 | FALSE | 0 | 0 | 1 | 1 | 2 | 0.50 | 0 | LINC02353 |
| rs28858724 | 4 | 0.03 | -0.34 | 0.17 | A | G | 0.04 | FALSE | 0 | 0 | 1 | 1 | 2 | 0.50 | 0 | LINC02353 |

leadSNP: Whether a given SNP is the locus lead SNP.

<tool>.CS: The posterior probability that a SNP is casual of the LiD phenotype.

Support: The number of fine-mapping tools that nominated the Consensus SNP

mean.PP: The mean SNP wise PP across fine mapping tools

mean.CS: If mean PP is greater than the 95% probability threshold (mean.PP > 0.95), then mean.CS is 1, else 0

**eTable 7.Colocalization hypotheses posterior probabilities**

| **ENSEMBL ID** | **HGNC** | **nsnps** | **PP.H0** | **PP.H1** | **PP.H2** | **PP.H3** | **PP.H4** | **ratio_PPH4_PPH3** |
| --- | --- | --- | --- | --- | --- | --- | --- | --- |
| ENSG00000162616 | DNAJB4 | 4840 | 6.76E-05 | 7.03E-05 | 0.23 | 0.24 | 0.52 | 2.17 |
| ENSG00000143067 | ZNF697 | 2881 | 2.60E-18 | 2.62E-19 | 0.72 | 0.07 | 0.21 | 2.93 |
| ENSG00000203782 | LORICRIN | 3572 | 5.25E-02 | 4.97E-03 | 0.74 | 0.07 | 0.14 | 1.96 |
| ENSG00000077254 | USP33 | 4552 | 4.54E-05 | 4.72E-05 | 0.43 | 0.44 | 0.13 | 0.30 |
| ENSG00000116266 | STXBP3 | 4434 | 1.07E-01 | 1.24E-02 | 0.72 | 0.08 | 0.08 | 0.92 |
| ENSG00000121940 | CLCC1 | 4311 | 7.44E-10 | 6.81E-11 | 0.85 | 0.08 | 0.07 | 0.92 |

nsnps: Number of overlapping SNPs between for each locus between the eqtl and the GWAS traits

PP.<hypothesis>: The posterior probability for each coloc hypothesis

ratio_PPH4_PPH3: The ratio of the H4 and H3 posterior porobabilities (ratio = H4/H3

**eTable 8. Candidate variants analysis.**

| **Nearest gene** | **SNP** | **MAF** | **BETA** | **SE** | **P-value** | **Direction** | **Publication** | **PMID** |
| --- | --- | --- | --- | --- | --- | --- | --- | --- |
| ANKK1 | rs1800497 | 0.21 | 0.24 | 0.09 | 8.89E-03 | +++-- | Rieck et al. 2012 | 23171335 |
| ANKK1 | rs2734849 | 0.50 | 0.18 | 0.08 | 2.11E-02 | +++++ | Rieck et al. 2012 | 23171335 |
| BDNF | rs6265 | 0.18 | 0.19 | 0.10 | 4.95E-02 | +++-+ | Foltynie et al. 2009  Kusters et al. 2018 | 18977816  29191473 |
| DRD2 | rs2283265 | 0.17 | 0.16 | 0.10 | 1.06E-01 | +++-- | Rieck et al. 2012 | 23171335 |
| DRD2 | rs6277 | 0.46 | 0.08 | 0.08 | 2.73E-01 | ----+ | Rieck et al. 2012 | 23171335 |
| DRD2 | rs1076560 | 0.17 | 0.15 | 0.10 | 1.42E-01 | +++-- | Rieck et al. 2012 | 23171335 |
| PRKCA | rs4790904 | 0.22 | -0.14 | 0.10 | 1.43E-01 | -++++ | Martin-Flores et al. 2018 | 29992529 |
| RPS6KB1 | rs1292034 | 0.42 | -0.13 | 0.08 | 1.08E-01 | ----- | Martin-Flores et al. 2018 | 29992529 |
| OPRM1 | rs1799971 | 0.12 | -0.13 | 0.12 | 3.04E-01 | +-+++ | Strong et al. 2006 | 16435402 |
| EIF4EBP2 | rs1043098 | 0.49 | 0.06 | 0.08 | 4.67E-01 | +-+-- | Martin-Flores et al. 2018 | 29992529 |
| SLC6A3 | rs393795 | 0.20 | 0.07 | 0.10 | 4.72E-01 | -++++ | Kaplan et al. 2014  Purcaro et al. 2018 | 24633632  30316985 |
| RICTOR | rs2043112 | 0.40 | 0.05 | 0.08 | 5.50E-01 | +++-+ | Martin-Flores et al. 2018 | 29992529 |
| **Nearest gene** | **SNP** | **MAF** | **BETA** | **SE** | **P-value** | **Direction** | **Publication** | **PMID** |
| HRAS | rs12628 | 0.35 | -0.04 | 0.08 | 5.89E-01 | +-++- | Martin-Flores et al. 2018 | 29992529 |
| RPS6KA2 | rs6456121 | 0.30 | 0.04 | 0.08 | 6.29E-01 | ++--+ | Martin-Flores et al. 2018 | 29992529 |
| COMT | rs4680 | 0.47 | -0.03 | 0.08 | 6.65E-01 | ++--- | Bialecka et al. 2004  de Lau et al. 2011  Hao et al. 2014  Cheshire al. 2014 | 15355491  24008922  25034874  22083803 |
| PRKN | rs1801582 | 0.16 | -0.04 | 0.11 | 7.01E-01 | -+-++ | Martin-Flores et al. 2018 | 29992529 |
| FCHSD1 | rs456998 | 0.49 | -0.03 | 0.08 | 7.17E-01 | +-+-+ | Martin-Flores et al. 2018 | 29992529 |
| DRD3 | rs6280 | 0.33 | 0.02 | 0.08 | 7.63E-01 | +-+-+ | Lee et al. 2011 | 20945430 |
| ADORA2A | rs3761422 | 0.37 | 0.02 | 0.08 | 7.71E-01 | +-++- | Rieck et al. 2015 | 25872644 |
| ADORA2A | rs2298383 | 0.40 | 0.02 | 0.08 | 8.39E-01 | -+-++ | Rieck et al. 2015 | 25872644 |
| HOMER1 | rs4704559 | 0.09 | -0.03 | 0.13 | 8.31E-01 | +++-- | Schumacher-Schuh et al. 2014 | 24126708 |

Direction: Indicates the directionality of the effect of the variant across substudies included on each study.

**eTable9. List of DJ-1, and PARKIN intergenic variants with a significance value lower than 0.05 on the raw GWAS meta-analysis**

| **SNP** | **CHR** | **BP** | **A1** | **A2** | **AF** | **HR** | **Beta** | **SE** | **P** | **N** | **GENE** |
| --- | --- | --- | --- | --- | --- | --- | --- | --- | --- | --- | --- |
| rs113276175 | 6 | 162600287 | a | g | 0.9827 | 0.22723 | -1.4818 | 0.395 | 0.00018 | 601 | DJ1 |
| rs75744512 | 6 | 162326363 | a | g | 0.0106 | 4.3991 | 1.4814 | 0.4411 | 0.00078 | 696 | DJ1 |
| rs144098700 | 6 | 162069891 | a | g | 0.9849 | 0.38612 | -0.9516 | 0.3047 | 0.00179 | 1708 | DJ1 |
| rs73782962 | 6 | 162075343 | a | g | 0.0159 | 2.32751 | 0.8448 | 0.2723 | 0.00192 | 1991 | DJ1 |
| rs111329397 | 6 | 162171336 | a | c | 0.0211 | 3.86013 | 1.3507 | 0.462 | 0.00346 | 1220 | DJ1 |
| rs113585246 | 6 | 162171423 | a | g | 0.0211 | 3.86013 | 1.3507 | 0.462 | 0.00346 | 1220 | DJ1 |
| rs112904254 | 6 | 162172584 | a | c | 0.0211 | 3.86013 | 1.3507 | 0.462 | 0.00346 | 1220 | DJ1 |
| rs73782959 | 6 | 162062632 | t | g | 0.0154 | 2.23468 | 0.8041 | 0.2813 | 0.00426 | 1991 | DJ1 |
| rs12111122 | 6 | 162068076 | a | t | 0.0154 | 2.23468 | 0.8041 | 0.2813 | 0.00426 | 1991 | DJ1 |
| rs57985302 | 6 | 162071287 | a | g | 0.0156 | 2.22888 | 0.8015 | 0.2813 | 0.00438 | 1991 | DJ1 |
| rs56403254 | 6 | 162071717 | a | g | 0.0156 | 2.22888 | 0.8015 | 0.2813 | 0.00438 | 1991 | DJ1 |
| rs143041505 | 6 | 162074039 | a | c | 0.9844 | 0.44866 | -0.8015 | 0.2813 | 0.00438 | 1991 | DJ1 |
| **SNP** | **CHR** | **BP** | **A1** | **A2** | **AF** | **HR** | **Beta** | **SE** | **P** | **N** | **GENE** |
| rs6912219 | 6 | 162074100 | a | c | 0.9844 | 0.44866 | -0.8015 | 0.2813 | 0.00438 | 1991 | DJ1 |
| rs189617732 | 6 | 162535958 | a | g | 0.9287 | 0.48384 | -0.726 | 0.256 | 0.00457 | 524 | DJ1 |
| rs6908330 | 6 | 162943061 | a | c | 0.8908 | 0.72044 | -0.3279 | 0.1164 | 0.00484 | 2687 | DJ1 |
| rs2846480 | 6 | 162946970 | t | c | 0.8908 | 0.72044 | -0.3279 | 0.1164 | 0.00484 | 2687 | DJ1 |
| rs2022998 | 6 | 162941823 | t | c | 0.88 | 0.7287 | -0.3165 | 0.1124 | 0.00486 | 2687 | DJ1 |
| rs2846482 | 6 | 162946152 | a | g | 0.1099 | 1.38472 | 0.3255 | 0.1164 | 0.00518 | 2687 | DJ1 |
| rs141726555 | 6 | 162250488 | t | c | 0.02 | 4.82109 | 1.573 | 0.5642 | 0.0053 | 241 | DJ1 |
| rs6455760 | 6 | 162110497 | a | g | 0.1245 | 1.3477 | 0.2984 | 0.1086 | 0.00598 | 2687 | DJ1 |
| rs182178663 | 6 | 162102507 | a | g | 0.0121 | 4.63947 | 1.5346 | 0.5599 | 0.00613 | 1014 | DJ1 |
| rs150977839 | 6 | 162076149 | t | c | 0.9847 | 0.44731 | -0.8045 | 0.2938 | 0.00617 | 1991 | DJ1 |
| rs9355901 | 6 | 161858094 | t | c | 0.2626 | 1.26226 | 0.2329 | 0.0853 | 0.00633 | 2687 | DJ1 |
| rs138643736 | 6 | 161825450 | t | c | 0.0126 | 44.2564 | 3.79 | 1.4047 | 0.00697 | 77 | DJ1 |
| rs76437736 | 6 | 162385389 | a | c | 0.9833 | 0.20458 | -1.5868 | 0.5941 | 0.00756 | 318 | DJ1 |
| rs147654033 | 6 | 162051481 | t | g | 0.0136 | 2.366 | 0.8612 | 0.3232 | 0.00771 | 1708 | DJ1 |
| **SNP** | **CHR** | **BP** | **A1** | **A2** | **AF** | **HR** | **Beta** | **SE** | **P** | **N** | **GENE** |
| rs113276175 | 6 | 162600287 | a | g | 0.9827 | 0.22723 | -1.4818 | 0.395 | 0.00018 | 601 | PARKN |
| rs75744512 | 6 | 162326363 | a | g | 0.0106 | 4.3991 | 1.4814 | 0.4411 | 0.00078 | 696 | PARKN |
| rs144098700 | 6 | 162069891 | a | g | 0.9849 | 0.38612 | -0.9516 | 0.3047 | 0.00179 | 1708 | PARKN |
| rs73782962 | 6 | 162075343 | a | g | 0.0159 | 2.32751 | 0.8448 | 0.2723 | 0.00192 | 1991 | PARKN |
| rs111329397 | 6 | 162171336 | a | c | 0.0211 | 3.86013 | 1.3507 | 0.462 | 0.00346 | 1220 | PARKN |
| rs113585246 | 6 | 162171423 | a | g | 0.0211 | 3.86013 | 1.3507 | 0.462 | 0.00346 | 1220 | PARKN |
| rs112904254 | 6 | 162172584 | a | c | 0.0211 | 3.86013 | 1.3507 | 0.462 | 0.00346 | 1220 | PARKN |
| rs73782959 | 6 | 162062632 | t | g | 0.0154 | 2.23468 | 0.8041 | 0.2813 | 0.00426 | 1991 | PARKN |
| rs12111122 | 6 | 162068076 | a | t | 0.0154 | 2.23468 | 0.8041 | 0.2813 | 0.00426 | 1991 | PARKN |
| rs57985302 | 6 | 162071287 | a | g | 0.0156 | 2.22888 | 0.8015 | 0.2813 | 0.00438 | 1991 | PARKN |
| rs56403254 | 6 | 162071717 | a | g | 0.0156 | 2.22888 | 0.8015 | 0.2813 | 0.00438 | 1991 | PARKN |
| rs143041505 | 6 | 162074039 | a | c | 0.9844 | 0.44866 | -0.8015 | 0.2813 | 0.00438 | 1991 | PARKN |
| rs6912219 | 6 | 162074100 | a | c | 0.9844 | 0.44866 | -0.8015 | 0.2813 | 0.00438 | 1991 | PARKN |
| rs189617732 | 6 | 162535958 | a | g | 0.9287 | 0.48384 | -0.726 | 0.256 | 0.00457 | 524 | PARKN |
| **SNP** | **CHR** | **BP** | **A1** | **A2** | **AF** | **HR** | **Beta** | **SE** | **P** | **N** | **GENE** |
| rs6908330 | 6 | 162943061 | a | c | 0.8908 | 0.72044 | -0.3279 | 0.1164 | 0.00484 | 2687 | PARKN |
| rs2846480 | 6 | 162946970 | t | c | 0.8908 | 0.72044 | -0.3279 | 0.1164 | 0.00484 | 2687 | PARKN |
| rs2022998 | 6 | 162941823 | t | c | 0.88 | 0.7287 | -0.3165 | 0.1124 | 0.00486 | 2687 | PARKN |
| rs2846482 | 6 | 162946152 | a | g | 0.1099 | 1.38472 | 0.3255 | 0.1164 | 0.00518 | 2687 | PARKN |
| rs141726555 | 6 | 162250488 | t | c | 0.02 | 4.82109 | 1.573 | 0.5642 | 0.0053 | 241 | PARKN |
| rs6455760 | 6 | 162110497 | a | g | 0.1245 | 1.3477 | 0.2984 | 0.1086 | 0.00598 | 2687 | PARKN |
| rs182178663 | 6 | 162102507 | a | g | 0.0121 | 4.63947 | 1.5346 | 0.5599 | 0.00613 | 1014 | PARKN |
| rs150977839 | 6 | 162076149 | t | c | 0.9847 | 0.44731 | -0.8045 | 0.2938 | 0.00617 | 1991 | PARKN |
| rs9355901 | 6 | 161858094 | t | c | 0.2626 | 1.26226 | 0.2329 | 0.0853 | 0.00633 | 2687 | PARKN |
| rs138643736 | 6 | 161825450 | t | c | 0.0126 | 44.2564 | 3.79 | 1.4047 | 0.00697 | 77 | PARKN |
| rs76437736 | 6 | 162385389 | a | c | 0.9833 | 0.20458 | -1.5868 | 0.5941 | 0.00756 | 318 | PARKN |
| rs147654033 | 6 | 162051481 | t | g | 0.0136 | 2.366 | 0.8612 | 0.3232 | 0.00771 | 1708 | PARKN |
| rs117765565 | 6 | 162871441 | a | t | 0.1003 | 0.64927 | -0.4319 | 0.1628 | 0.00797 | 2445 | PARKN |
| rs1801474 | 6 | 162622197 | t | c | 0.0146 | 2.19614 | 0.7867 | 0.297 | 0.00808 | 2687 | PARKN |
| **SNP** | **CHR** | **BP** | **A1** | **A2** | **AF** | **HR** | **Beta** | **SE** | **P** | **N** | **GENE** |
| rs9458505 | 6 | 162725665 | t | c | 0.0913 | 1.42333 | 0.353 | 0.1339 | 0.00838 | 2404 | PARKN |
| rs6914057 | 6 | 162871262 | t | c | 0.9005 | 1.4981 | 0.4042 | 0.1538 | 0.00857 | 2687 | PARKN |
| rs12662364 | 6 | 161855965 | a | g | 0.2642 | 1.24995 | 0.2231 | 0.085 | 0.00867 | 2687 | PARKN |
| rs62437986 | 6 | 162582554 | t | c | 0.986 | 0.2644 | -1.3303 | 0.5092 | 0.00899 | 1297 | PARKN |
| rs55912218 | 6 | 162307775 | t | taa | 0.7153 | 1.86526 | 0.6234 | 0.2409 | 0.00966 | 281 | PARKN |
| rs149634732 | 6 | 162047729 | t | c | 0.0136 | 2.10244 | 0.7431 | 0.2872 | 0.00968 | 1991 | PARKN |
| rs9347542 | 6 | 162235691 | t | c | 0.4193 | 1.46594 | 0.3825 | 0.1486 | 0.01006 | 524 | PARKN |
| rs151243521 | 6 | 162460413 | t | c | 0.9833 | 0.2195 | -1.5164 | 0.5893 | 0.01007 | 318 | PARKN |
| rs112858840 | 6 | 163137317 | c | g | 0.9833 | 0.4205 | -0.8663 | 0.342 | 0.0113 | 1220 | PARKN |
| rs57374961 | 6 | 162615335 | c | g | 0.0151 | 2.09552 | 0.7398 | 0.2931 | 0.0116 | 2687 | PARKN |
| rs78718632 | 6 | 162877658 | a | g | 0.1009 | 0.68537 | -0.3778 | 0.1514 | 0.01258 | 2687 | PARKN |
| rs74475107 | 6 | 162877889 | a | g | 0.8991 | 1.45907 | 0.3778 | 0.1514 | 0.01258 | 2687 | PARKN |
| rs143941834 | 6 | 162640507 | a | g | 0.9842 | 0.29724 | -1.2132 | 0.4873 | 0.01278 | 524 | PARKN |
| rs75342128 | 6 | 162652000 | a | g | 0.9842 | 0.29724 | -1.2132 | 0.4873 | 0.01278 | 524 | PARKN |
| **SNP** | **CHR** | **BP** | **A1** | **A2** | **AF** | **HR** | **Beta** | **SE** | **P** | **N** | **GENE** |
| rs76146820 | 6 | 162707685 | a | g | 0.0158 | 3.36423 | 1.2132 | 0.4873 | 0.01278 | 524 | PARKN |
| rs9365407 | 6 | 162785031 | a | g | 0.8226 | 0.78718 | -0.2393 | 0.0964 | 0.01305 | 2687 | PARKN |
| rs6941157 | 6 | 162936188 | t | c | 0.0565 | 1.44672 | 0.3693 | 0.1488 | 0.01307 | 2687 | PARKN |
| rs12214138 | 6 | 162107046 | t | c | 0.9638 | 0.64076 | -0.4451 | 0.1829 | 0.01494 | 2687 | PARKN |
| rs118180252 | 6 | 162097162 | t | c | 0.0233 | 1.65367 | 0.503 | 0.2073 | 0.01524 | 2687 | PARKN |
| rs60095244 | 6 | 162946561 | t | c | 0.0564 | 1.42975 | 0.3575 | 0.1489 | 0.01636 | 2687 | PARKN |
| rs60074916 | 6 | 162946564 | t | c | 0.0564 | 1.42975 | 0.3575 | 0.1489 | 0.01636 | 2687 | PARKN |
| rs16892937 | 6 | 162125691 | t | c | 0.0188 | 1.74072 | 0.5543 | 0.2312 | 0.01651 | 2687 | PARKN |
| rs192576507 | 6 | 162715710 | t | c | 0.9916 | 0.02812 | -3.5714 | 1.4967 | 0.01703 | 77 | PARKN |
| rs80330859 | 6 | 162859951 | c | g | 0.9031 | 1.44037 | 0.3649 | 0.1533 | 0.01726 | 2687 | PARKN |
| rs59996420 | 6 | 162122039 | a | g | 0.9811 | 0.57782 | -0.5485 | 0.2312 | 0.01767 | 2687 | PARKN |
| rs62430696 | 6 | 162752612 | t | c | 0.9808 | 0.56666 | -0.568 | 0.24 | 0.01795 | 2687 | PARKN |
| rs62430697 | 6 | 162753224 | t | c | 0.0192 | 1.76473 | 0.568 | 0.24 | 0.01795 | 2687 | PARKN |
| rs9458572 | 6 | 162935431 | t | c | 0.0507 | 1.44023 | 0.3648 | 0.1543 | 0.0181 | 2687 | PARKN |
| **SNP** | **CHR** | **BP** | **A1** | **A2** | **AF** | **HR** | **Beta** | **SE** | **P** | **N** | **GENE** |
| rs374239029 | 6 | 162777567 | a | at | 0.7438 | 1.87142 | 0.6267 | 0.2653 | 0.01816 | 283 | PARKN |
| rs73597197 | 6 | 162124704 | t | c | 0.9808 | 0.58077 | -0.5434 | 0.2313 | 0.01882 | 2687 | PARKN |
| rs9365410 | 6 | 162795058 | t | c | 0.061 | 1.40649 | 0.3411 | 0.1462 | 0.0196 | 2687 | PARKN |
| rs79565809 | 6 | 162119637 | a | g | 0.0189 | 1.71412 | 0.5389 | 0.2312 | 0.01974 | 2687 | PARKN |
| rs1016085 | 6 | 162242404 | t | c | 0.7634 | 0.81595 | -0.2034 | 0.0873 | 0.01979 | 2687 | PARKN |
| rs116357950 | 6 | 161970866 | t | c | 0.0338 | 1.64001 | 0.4947 | 0.2125 | 0.01992 | 2687 | PARKN |
| rs186614596 | 6 | 162534882 | t | c | 0.0589 | 0.25576 | -1.3635 | 0.5862 | 0.02001 | 524 | PARKN |
| rs111367465 | 6 | 162535731 | a | c | 0.0589 | 0.25576 | -1.3635 | 0.5862 | 0.02001 | 524 | PARKN |
| rs62437978 | 6 | 162573952 | a | g | 0.0084 | 29.45312 | 3.3828 | 1.4632 | 0.02078 | 77 | PARKN |
| rs140706721 | 6 | 162477163 | t | c | 0.0085 | 3.42123 | 1.23 | 0.5357 | 0.02166 | 773 | PARKN |
| rs146099223 | 6 | 162563818 | a | g | 0.0085 | 3.42123 | 1.23 | 0.5357 | 0.02166 | 773 | PARKN |
| rs150553548 | 6 | 162695552 | a | t | 0.0104 | 2.66153 | 0.9789 | 0.4264 | 0.02169 | 1390 | PARKN |
| rs144698234 | 6 | 162715035 | a | g | 0.0104 | 2.66153 | 0.9789 | 0.4264 | 0.02169 | 1390 | PARKN |
| rs79026473 | 6 | 162097898 | a | c | 0.9761 | 0.62744 | -0.4661 | 0.2033 | 0.02185 | 2687 | PARKN |
| **SNP** | **CHR** | **BP** | **A1** | **A2** | **AF** | **HR** | **Beta** | **SE** | **P** | **N** | **GENE** |
| rs9458348 | 6 | 162149539 | t | c | 0.7107 | 0.82456 | -0.1929 | 0.0841 | 0.02186 | 2687 | PARKN |
| rs74922818 | 6 | 162696500 | a | c | 0.0107 | 2.65727 | 0.9773 | 0.4264 | 0.02191 | 1390 | PARKN |
| rs138089003 | 6 | 162354879 | t | g | 0.0168 | 7.86797 | 2.0628 | 0.9004 | 0.02197 | 77 | PARKN |
| rs138771484 | 6 | 162432979 | t | c | 0.979 | 0.1271 | -2.0628 | 0.9004 | 0.02197 | 77 | PARKN |
| rs60883055 | 6 | 162611302 | t | c | 0.9769 | 0.57132 | -0.5598 | 0.2451 | 0.02238 | 2687 | PARKN |
| rs140432232 | 6 | 162843810 | c | g | 0.9028 | 1.41397 | 0.3464 | 0.1519 | 0.0226 | 2687 | PARKN |
| rs192586459 | 6 | 162876746 | t | c | 0.1081 | 0.45841 | -0.78 | 0.3422 | 0.02263 | 524 | PARKN |
| rs188082227 | 6 | 162723904 | a | t | 0.0168 | 10.47614 | 2.3491 | 1.033 | 0.02296 | 77 | PARKN |
| rs6930628 | 6 | 162110806 | a | g | 0.0218 | 1.65004 | 0.5008 | 0.2203 | 0.02301 | 2687 | PARKN |
| rs16892913 | 6 | 162112309 | a | t | 0.9782 | 0.60605 | -0.5008 | 0.2203 | 0.02301 | 2687 | PARKN |
| rs76290633 | 6 | 162116665 | a | g | 0.0218 | 1.65004 | 0.5008 | 0.2203 | 0.02301 | 2687 | PARKN |
| rs4709541 | 6 | 162103294 | a | g | 0.2062 | 0.78852 | -0.2376 | 0.1046 | 0.02306 | 2687 | PARKN |
| rs11751911 | 6 | 162235611 | t | c | 0.2516 | 0.80961 | -0.2112 | 0.0931 | 0.02329 | 2687 | PARKN |
| rs4708931 | 6 | 162190804 | a | c | 0.949 | 0.65423 | -0.4243 | 0.188 | 0.02402 | 1914 | PARKN |
| **SNP** | **CHR** | **BP** | **A1** | **A2** | **AF** | **HR** | **Beta** | **SE** | **P** | **N** | **GENE** |
| rs4455643 | 6 | 161852968 | t | c | 0.7346 | 0.82572 | -0.1915 | 0.085 | 0.02437 | 2687 | PARKN |
| rs9347509 | 6 | 161853203 | a | g | 0.7346 | 0.82572 | -0.1915 | 0.085 | 0.02437 | 2687 | PARKN |
| rs62436001 | 6 | 162391014 | t | c | 0.0159 | 2.99936 | 1.0984 | 0.4884 | 0.0245 | 1220 | PARKN |
| rs77460805 | 6 | 162773895 | c | g | 0.892 | 1.37479 | 0.3183 | 0.1423 | 0.02535 | 2687 | PARKN |
| rs10755584 | 6 | 162469736 | t | c | 0.8648 | 0.77942 | -0.2492 | 0.1115 | 0.02542 | 2687 | PARKN |
| rs117230217 | 6 | 162296735 | t | c | 0.0283 | 2.7871 | 1.025 | 0.4627 | 0.02672 | 524 | PARKN |
| rs9295181 | 6 | 162481460 | t | c | 0.5783 | 0.83937 | -0.1751 | 0.0793 | 0.02729 | 2687 | PARKN |
| rs568459658 | 6 | 162192301 | t | c | 0.9833 | 0.54313 | -0.6104 | 0.2772 | 0.02765 | 2687 | PARKN |
| rs557347438 | 6 | 162227423 | a | c | 0.0168 | 22.20017 | 3.1001 | 1.412 | 0.02812 | 77 | PARKN |
| rs62429614 | 6 | 162940960 | t | c | 0.9816 | 0.5351 | -0.6253 | 0.2871 | 0.0294 | 2610 | PARKN |
| rs78907851 | 6 | 162727657 | t | c | 0.0582 | 1.38085 | 0.3227 | 0.1486 | 0.02984 | 2687 | PARKN |
| rs9364650 | 6 | 162728256 | a | c | 0.9418 | 0.72419 | -0.3227 | 0.1486 | 0.02984 | 2687 | PARKN |
| rs75157574 | 6 | 162434018 | t | c | 0.025 | 1.7291 | 0.5476 | 0.2541 | 0.03112 | 1991 | PARKN |
| rs79220797 | 6 | 162835227 | a | g | 0.9026 | 1.38292 | 0.3242 | 0.1506 | 0.03133 | 2687 | PARKN |
| **SNP** | **CHR** | **BP** | **A1** | **A2** | **AF** | **HR** | **Beta** | **SE** | **P** | **N** | **GENE** |
| rs10945811 | 6 | 162738512 | a | g | 0.0582 | 1.37754 | 0.3203 | 0.1488 | 0.03134 | 2687 | PARKN |
| rs9355898 | 6 | 161847010 | t | c | 0.2694 | 0.67086 | -0.3992 | 0.1858 | 0.03164 | 524 | PARKN |
| rs75923621 | 6 | 162830552 | a | t | 0.097 | 0.72368 | -0.3234 | 0.1506 | 0.03173 | 2687 | PARKN |
| rs79699210 | 6 | 162834650 | a | g | 0.097 | 0.72368 | -0.3234 | 0.1506 | 0.03173 | 2687 | PARKN |
| rs74892485 | 6 | 162834786 | t | c | 0.097 | 0.72368 | -0.3234 | 0.1506 | 0.03173 | 2687 | PARKN |
| rs78174510 | 6 | 162839960 | t | c | 0.9026 | 1.38182 | 0.3234 | 0.1506 | 0.03175 | 2687 | PARKN |
| rs12210160 | 6 | 161814907 | a | g | 0.897 | 1.34299 | 0.2949 | 0.1374 | 0.0318 | 2687 | PARKN |
| rs2186815 | 6 | 162886686 | a | g | 0.052 | 1.40214 | 0.338 | 0.1575 | 0.03186 | 2687 | PARKN |
| rs78964633 | 6 | 162837366 | t | c | 0.0972 | 0.7239 | -0.3231 | 0.1506 | 0.03193 | 2687 | PARKN |
| rs11753929 | 6 | 162088198 | t | c | 0.9785 | 0.59156 | -0.525 | 0.245 | 0.03211 | 2687 | PARKN |
| rs71653628 | 6 | 161770811 | t | g | 0.044 | 1.44773 | 0.37 | 0.1727 | 0.03219 | 2687 | PARKN |
| rs146512832 | 6 | 162220484 | c | g | 0.9876 | 0.25173 | -1.3794 | 0.6462 | 0.0328 | 283 | PARKN |
| rs62430699 | 6 | 162774531 | a | g | 0.01 | 5.76036 | 1.751 | 0.8253 | 0.03387 | 241 | PARKN |
| rs747295 | 6 | 162888355 | t | c | 0.9061 | 0.77136 | -0.2596 | 0.1227 | 0.03445 | 2687 | PARKN |
| **SNP** | **CHR** | **BP** | **A1** | **A2** | **AF** | **HR** | **Beta** | **SE** | **P** | **N** | **GENE** |
| rs75888372 | 6 | 162804109 | t | g | 0.9029 | 1.37383 | 0.3176 | 0.1505 | 0.03485 | 2687 | PARKN |
| rs114696305 | 6 | 162812684 | a | g | 0.0974 | 0.72826 | -0.3171 | 0.1504 | 0.03494 | 2687 | PARKN |
| rs35029799 | 6 | 162283430 | a | g | 0.9207 | 0.76208 | -0.2717 | 0.1294 | 0.03577 | 2687 | PARKN |
| rs6455759 | 6 | 162106439 | t | c | 0.2119 | 0.80541 | -0.2164 | 0.1031 | 0.03581 | 2687 | PARKN |
| rs542536086 | 6 | 163141639 | t | c | 0.0117 | 4.93573 | 1.5965 | 0.7635 | 0.03653 | 241 | PARKN |
| rs78648977 | 6 | 162803458 | t | g | 0.01 | 5.47504 | 1.7002 | 0.8151 | 0.03699 | 241 | PARKN |
| rs9347599 | 6 | 162611892 | t | c | 0.0145 | 1.83639 | 0.6078 | 0.2915 | 0.03707 | 2687 | PARKN |
| rs9355933 | 6 | 162190430 | a | t | 0.0505 | 1.48676 | 0.3966 | 0.1904 | 0.03719 | 1914 | PARKN |
| rs4708930 | 6 | 162190759 | t | c | 0.0505 | 1.48676 | 0.3966 | 0.1904 | 0.03719 | 1914 | PARKN |
| rs1001091 | 6 | 162237057 | a | g | 0.7275 | 1.20889 | 0.1897 | 0.091 | 0.03719 | 2687 | PARKN |
| rs2849564 | 6 | 162459664 | t | c | 0.3709 | 1.17998 | 0.1655 | 0.0794 | 0.03728 | 2687 | PARKN |
| rs79084151 | 6 | 162832167 | t | g | 0.8978 | 1.35256 | 0.302 | 0.1452 | 0.03758 | 2687 | PARKN |
| rs1122327 | 6 | 162235108 | t | c | 0.3028 | 1.1891 | 0.1732 | 0.0833 | 0.0376 | 2687 | PARKN |
| rs74778695 | 6 | 162931570 | a | g | 0.0229 | 2.35608 | 0.857 | 0.4123 | 0.03768 | 601 | PARKN |
| **SNP** | **CHR** | **BP** | **A1** | **A2** | **AF** | **HR** | **Beta** | **SE** | **P** | **N** | **GENE** |
| rs6914080 | 6 | 162871297 | t | c | 0.6423 | 0.84764 | -0.1653 | 0.0795 | 0.03769 | 2687 | PARKN |
| rs9295150 | 6 | 161885096 | a | g | 0.0213 | 1.64132 | 0.4955 | 0.2386 | 0.03781 | 2610 | PARKN |
| rs9458453 | 6 | 162502852 | a | t | 0.5467 | 1.17774 | 0.1636 | 0.0788 | 0.03794 | 2687 | PARKN |
| rs200316663 | 6 | 162697733 | g | gtgtc | 0.9466 | 2.68451 | 0.9875 | 0.4759 | 0.03797 | 524 | PARKN |
| rs192572757 | 6 | 162531271 | t | c | 0.979 | 0.09673 | -2.3358 | 1.1288 | 0.03851 | 77 | PARKN |
| rs141512658 | 6 | 162552268 | a | c | 0.021 | 10.33773 | 2.3358 | 1.1288 | 0.03851 | 77 | PARKN |
| rs4314474 | 6 | 162518553 | t | c | 0.3262 | 1.39766 | 0.3348 | 0.162 | 0.03869 | 524 | PARKN |
| rs75856612 | 6 | 162513392 | a | g | 0.9883 | 0.19311 | -1.6445 | 0.7973 | 0.03916 | 241 | PARKN |
| rs73013498 | 6 | 162139284 | a | g | 0.2006 | 1.21933 | 0.1983 | 0.0963 | 0.0395 | 2687 | PARKN |
| rs2851401 | 6 | 162459762 | t | c | 0.4967 | 1.1721 | 0.1588 | 0.0772 | 0.03984 | 2687 | PARKN |
| rs7758666 | 6 | 162249776 | a | c | 0.2845 | 1.19017 | 0.1741 | 0.0848 | 0.04005 | 2687 | PARKN |
| rs12205900 | 6 | 162094674 | a | g | 0.0339 | 1.4755 | 0.389 | 0.1901 | 0.04071 | 2687 | PARKN |
| rs60798458 | 6 | 161847112 | a | c | 0.1307 | 1.7369 | 0.5521 | 0.2705 | 0.04126 | 283 | PARKN |
| rs9365409 | 6 | 162792379 | t | c | 0.1907 | 1.20852 | 0.1894 | 0.0931 | 0.04194 | 2687 | PARKN |
| **SNP** | **CHR** | **BP** | **A1** | **A2** | **AF** | **HR** | **Beta** | **SE** | **P** | **N** | **GENE** |
| rs117076984 | 6 | 162991852 | c | g | 0.0302 | 1.56721 | 0.4493 | 0.2215 | 0.04248 | 2687 | PARKN |
| rs112765480 | 6 | 162770514 | c | g | 0.9805 | 0.60854 | -0.4967 | 0.2459 | 0.04342 | 2687 | PARKN |
| rs12193568 | 6 | 162557719 | a | g | 0.7507 | 0.83644 | -0.1786 | 0.0886 | 0.04393 | 2687 | PARKN |
| rs4709569 | 6 | 162503148 | a | c | 0.5455 | 1.17128 | 0.1581 | 0.0787 | 0.04455 | 2687 | PARKN |
| rs1001090 | 6 | 162237100 | a | g | 0.7292 | 1.19973 | 0.1821 | 0.0909 | 0.04516 | 2687 | PARKN |
| rs9458452 | 6 | 162501019 | t | c | 0.5448 | 1.17 | 0.157 | 0.0788 | 0.04621 | 2687 | PARKN |
| rs13217906 | 6 | 161804399 | a | g | 0.1024 | 0.76231 | -0.2714 | 0.1363 | 0.04647 | 2687 | PARKN |
| rs10806755 | 6 | 162495315 | a | g | 0.2234 | 1.41468 | 0.3469 | 0.1745 | 0.04677 | 524 | PARKN |
| rs78600213 | 6 | 161939935 | c | g | 0.0161 | 1.74334 | 0.5558 | 0.2796 | 0.04686 | 2687 | PARKN |
| rs1893545 | 6 | 162451368 | t | g | 0.2005 | 1.21046 | 0.191 | 0.0966 | 0.04804 | 2687 | PARKN |
| rs79948313 | 6 | 162725598 | a | t | 0.0592 | 1.33442 | 0.2885 | 0.1462 | 0.04845 | 2687 | PARKN |
| rs138485300 | 6 | 162463415 | a | c | 0.0097 | 2.19416 | 0.7858 | 0.3983 | 0.0485 | 2163 | PARKN |
| rs12213394 | 6 | 161797681 | a | g | 0.1021 | 0.76414 | -0.269 | 0.1364 | 0.04859 | 2687 | PARKN |
| rs12213397 | 6 | 161797690 | t | g | 0.1021 | 0.76414 | -0.269 | 0.1364 | 0.04859 | 2687 | PARKN |
| **SNP** | **CHR** | **BP** | **A1** | **A2** | **AF** | **HR** | **Beta** | **SE** | **P** | **N** | **GENE** |
| rs10806754 | 6 | 162495290 | t | g | 0.7747 | 0.70957 | -0.3431 | 0.1741 | 0.04877 | 524 | PARKN |
| rs117406278 | 6 | 162431177 | t | c | 0.9295 | 0.64025 | -0.4459 | 0.2267 | 0.04914 | 524 | PARKN |
| rs13191078 | 6 | 162476705 | a | c | 0.4528 | 0.85607 | -0.1554 | 0.079 | 0.04927 | 2687 | PARKN |
| rs1018462 | 6 | 162141896 | a | g | 0.6623 | 0.85266 | -0.1594 | 0.0811 | 0.04945 | 2687 | PARKN |
| rs146701725 | 6 | 162725858 | c | g | 0.0142 | 3.35483 | 1.2104 | 0.6161 | 0.04945 | 282 | PARKN |
| rs2849565 | 6 | 162452162 | a | g | 0.1475 | 1.23937 | 0.2146 | 0.1093 | 0.04947 | 2687 | PARKN |
| rs139103004 | 6 | 162416907 | t | c | 0.0138 | 2.18475 | 0.7815 | 0.3978 | 0.04949 | 1014 | PARKN |
| rs9458252 | 6 | 161860169 | t | c | 0.5577 | 0.8559 | -0.1556 | 0.0795 | 0.05029 | 2687 | PARKN |

**eTable 10. Table of SNPs used to derive the GRS in the TPD cohort**

| **SNP** | **CHR** | **BP** | **MAF** | **BETA** | **SE** | **P-value** |
| --- | --- | --- | --- | --- | --- | --- |
| 4:32435284 | 4 | 32435284 | 0.0194 | 1.1245 | 0.1866 | 1.673e-09 |
| 16:17044975 | 16 | 17044975 | 0.9764 | -1.14 | 0.1962 | 6.265e-09 |
| 1:53778300 | 1 | 53778300 | 0.0233 | 1.0163 | 0.1796 | 1.527e-08 |
| 1:168645690 | 1 | 168645690 | 0.0466 | 0.7671 | 0.1423 | 7.037e-08 |
| 1:80950480 | 1 | 80950480 | 0.9548 | -0.7594 | 0.1419 | 8.692e-08 |
| 9:22664277 | 9 | 22664277 | 0.0143 | 1.2588 | 0.2356 | 9.192e-08 |
| 2:47299166 | 2 | 47299166 | 0.019 | 1.0791 | 0.205 | 1.416e-07 |
| 13:73750127 | 13 | 73750127 | 0.0672 | 0.6937 | 0.1343 | 2.41e-07 |
| 3:8537619 | 3 | 8537619 | 0.9334 | -0.6374 | 0.1256 | 3.856e-07 |
| 3:4714509 | 3 | 4714509 | 0.0287 | 0.8517 | 0.1693 | 4.901e-07 |
| 2:242660630 | 2 | 242660630 | 0.0144 | 1.237 | 0.2459 | 4.905e-07 |
| 12:48124937 | 12 | 48124937 | 0.0187 | 1.0675 | 0.2124 | 4.989e-07 |
| 5:108270767 | 5 | 108270767 | 0.0137 | 1.2506 | 0.2509 | 6.221e-07 |
| **SNP** | **CHR** | **BP** | **MAF** | **BETA** | **SE** | **P-value** |
| 8:138767782 | 8 | 138767782 | 0.0348 | 0.8365 | 0.168 | 6.423e-07 |
| 14:22023873 | 14 | 22023873 | 0.9868 | -1.1512 | 0.2321 | 7.04e-07 |
| 7:46390116 | 7 | 46390116 | 0.0224 | 0.9524 | 0.1936 | 8.679e-07 |
| 20:51996645 | 20 | 51996645 | 0.0139 | 1.0939 | 0.2227 | 9.062e-07 |
| 2:176515033 | 2 | 176515033 | 0.0596 | 0.6755 | 0.1376 | 9.191e-07 |
| 4:41614236 | 4 | 41614236 | 0.9878 | -1.2227 | 0.2496 | 9.637e-07 |
| 7:125755287 | 7 | 125755287 | 0.0321 | 0.881 | 0.1801 | 9.986e-07 |
| 11:93486449 | 11 | 93486449 | 0.9842 | -1.2365 | 0.2533 | 1.055e-06 |
| 19:30990206 | 19 | 30990206 | 0.9762 | -1.0323 | 0.2119 | 1.104e-06 |
| 6:16636328 | 6 | 16636328 | 0.0405 | 0.7637 | 0.157 | 1.152e-06 |
| 3:124888091 | 3 | 124888091 | 0.9463 | -0.6963 | 0.1432 | 1.166e-06 |
| 1:81453727 | 1 | 81453727 | 0.9875 | -1.1372 | 0.2342 | 1.204e-06 |
| 19:954677 | 19 | 954677 | 0.0266 | 0.9056 | 0.1867 | 1.233e-06 |
| 16:78198192 | 16 | 78198192 | 0.2731 | 0.4042 | 0.0835 | 1.287e-06 |
| **SNP** | **CHR** | **BP** | **MAF** | **BETA** | **SE** | **P-value** |
| 4:64494944 | 4 | 64494944 | 0.0206 | 0.9275 | 0.1923 | 1.414e-06 |
| 5:132249144 | 5 | 132249144 | 0.016 | 1.0617 | 0.2204 | 1.462e-06 |
| 2:62427404 | 2 | 62427404 | 0.0191 | 0.9786 | 0.2032 | 1.463e-06 |
| 15:61097179 | 15 | 61097179 | 0.0174 | 1.4453 | 0.3005 | 1.514e-06 |
| 8:103719246 | 8 | 103719246 | 0.0113 | 1.3437 | 0.2804 | 1.656e-06 |
| 3:128349376 | 3 | 128349376 | 0.9599 | -0.7262 | 0.1516 | 1.672e-06 |
| 1:106719668 | 1 | 106719668 | 0.0302 | 0.8321 | 0.1739 | 1.707e-06 |
| 7:157406470 | 7 | 157406470 | 0.9203 | -0.5601 | 0.1171 | 1.733e-06 |
| 9:124098846 | 9 | 124098846 | 0.0199 | 0.9737 | 0.2047 | 1.972e-06 |
| 18:1941742 | 18 | 1941742 | 0.0169 | 1.039 | 0.2187 | 2.035e-06 |
| 5:75719143 | 5 | 75719143 | 0.0156 | 1.2049 | 0.2538 | 2.052e-06 |
| 13:29849305 | 13 | 29849305 | 0.023 | 0.8936 | 0.1883 | 2.084e-06 |
| 13:95085417 | 13 | 95085417 | 0.1269 | 0.4899 | 0.1035 | 2.204e-06 |
| 4:94768877 | 4 | 94768877 | 0.0118 | 1.1815 | 0.2501 | 2.317e-06 |
| **SNP** | **CHR** | **BP** | **MAF** | **BETA** | **SE** | **P-value** |
| 18:54853305 | 18 | 54853305 | 0.023 | 0.9298 | 0.1977 | 2.57e-06 |
| 6:115767948 | 6 | 115767948 | 0.0256 | 0.8527 | 0.1819 | 2.765e-06 |
| 6:118528190 | 6 | 118528190 | 0.0334 | 0.8212 | 0.1756 | 2.932e-06 |
| 13:110061713 | 13 | 110061713 | 0.1381 | 0.4664 | 0.0998 | 2.999e-06 |
| 16:686398 | 16 | 686398 | 0.9824 | -1.0484 | 0.2247 | 3.081e-06 |
| 12:131890366 | 12 | 131890366 | 0.0111 | 1.2191 | 0.2621 | 3.301e-06 |
| 5:134774168 | 5 | 134774168 | 0.0111 | 1.4659 | 0.3159 | 3.49e-06 |
| 4:31553640 | 4 | 31553640 | 0.9863 | -1.1423 | 0.2463 | 3.531e-06 |
| 3:5287334 | 3 | 5287334 | 0.9855 | -1.1169 | 0.2411 | 3.61e-06 |
| 7:157515114 | 7 | 157515114 | 0.0131 | 1.0623 | 0.2294 | 3.629e-06 |
| 18:42296688 | 18 | 42296688 | 0.0127 | 1.216 | 0.2627 | 3.662e-06 |
| 2:37411014 | 2 | 37411014 | 0.9597 | -0.715 | 0.1544 | 3.662e-06 |
| 3:73128332 | 3 | 73128332 | 0.9876 | -1.2229 | 0.2643 | 3.728e-06 |
| 11:4852271 | 11 | 4852271 | 0.0892 | 0.5495 | 0.119 | 3.902e-06 |
| **SNP** | **CHR** | **BP** | **MAF** | **BETA** | **SE** | **P-value** |
| 11:99250689 | 11 | 99250689 | 0.0193 | 0.9499 | 0.2058 | 3.906e-06 |
| 13:95774697 | 13 | 95774697 | 0.9329 | -0.5648 | 0.1224 | 3.952e-06 |
| 1:78130243 | 1 | 78130243 | 0.2654 | 0.3958 | 0.0859 | 4.034e-06 |
| 1:71964730 | 1 | 71964730 | 0.9825 | -0.9834 | 0.2135 | 4.113e-06 |
| 16:20409582 | 16 | 20409582 | 0.9864 | -1.1181 | 0.2432 | 4.268e-06 |
| 9:81601411 | 9 | 81601411 | 0.0207 | 1.047 | 0.2278 | 4.291e-06 |
| 11:6636154 | 11 | 6636154 | 0.0133 | 1.2502 | 0.2722 | 4.364e-06 |
| 20:6363480 | 20 | 6363480 | 0.3078 | 0.3717 | 0.081 | 4.494e-06 |
| 14:59208732 | 14 | 59208732 | 0.1591 | 0.454 | 0.099 | 4.529e-06 |
| 8:135945999 | 8 | 135945999 | 0.9825 | -1.037 | 0.2266 | 4.732e-06 |
| 3:82436301 | 3 | 82436301 | 0.0108 | 1.3701 | 0.2997 | 4.853e-06 |
| 1:55014822 | 1 | 55014822 | 0.9634 | -0.7579 | 0.1659 | 4.945e-06 |

**eTable 11. MoCa and UPDRS score comparison between PD-LiD and PD groups**

| **Variable** | **method** | **p.value** | **statistic** | **PD group**  **mean(sd)** | **PD-LID group**  **mean(sd)** |
| --- | --- | --- | --- | --- | --- |
| moca_bl | Wilcoxon rank sum test with continuity correction | 2.1e-05 | 73754.5 | 25.16(3.31) | 26.09(3.56) |
| moca_visit | Wilcoxon rank sum test with continuity correction | 0.01 | 77249.5 | 24.34(4.77) | 25.35(4.11) |
| updrs_III_bl | Welch Two Sample t-test | 0.15 | -1.45 | 22.2(11.6) | 23.72(12.1) |
| updrs_III_visit | Welch Two Sample t-test | 0.25 | 1.15 | 31.91(16.7) | 30.42(14.0) |

moca_bl = Moca average scores for the LiD and PD group at baseline

moca_visit = Moca average scores for the LiD and PD group at the time LiD was developed or at the last visit available

updrsIII_bl = MDS-UPDRS III averages scores for the LiD and PD group at baseline

Updrs_iii_visit = MDS-UPDRS III average scores for the LiD and PD group at the time LiD was developed or at the last visit available

**3. eFigures.**

**eFigure 1. Quality control flowchart**

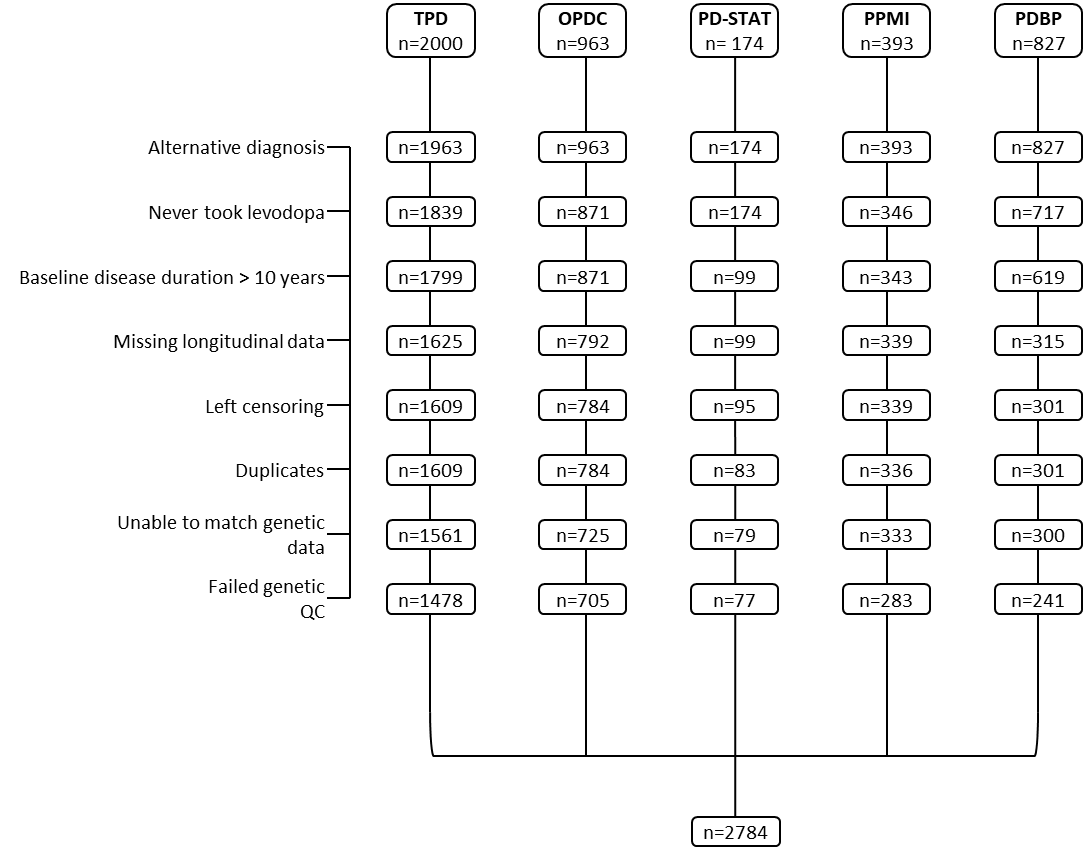

**eFigure 2. LiD risk factors Kaplan Meyer curves.** Kaplan-Meier curve for Survival probability (LID free probability) based on gender (A), age at onset (AAO) (B) baseline body mass index (BMI) (C), and smoking status (D). The P-value (P) showing the significance of differences on the survival probability is given on each plot. Number at risk represents the number of PD patients remaining on the study at the different time points (0, 5, 10, 15 years). The colour expansion on each curve represents the confidence interval (CI).

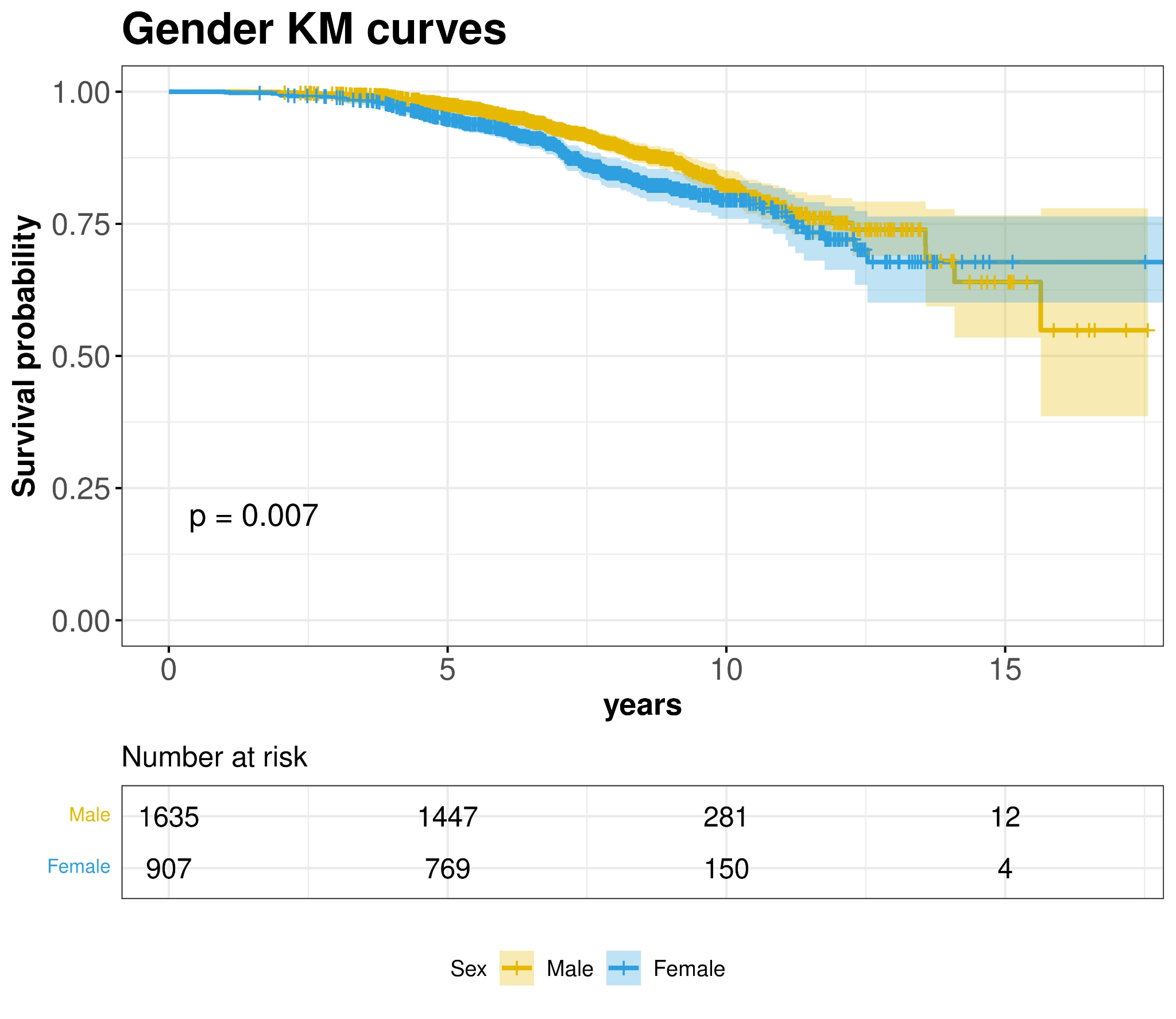

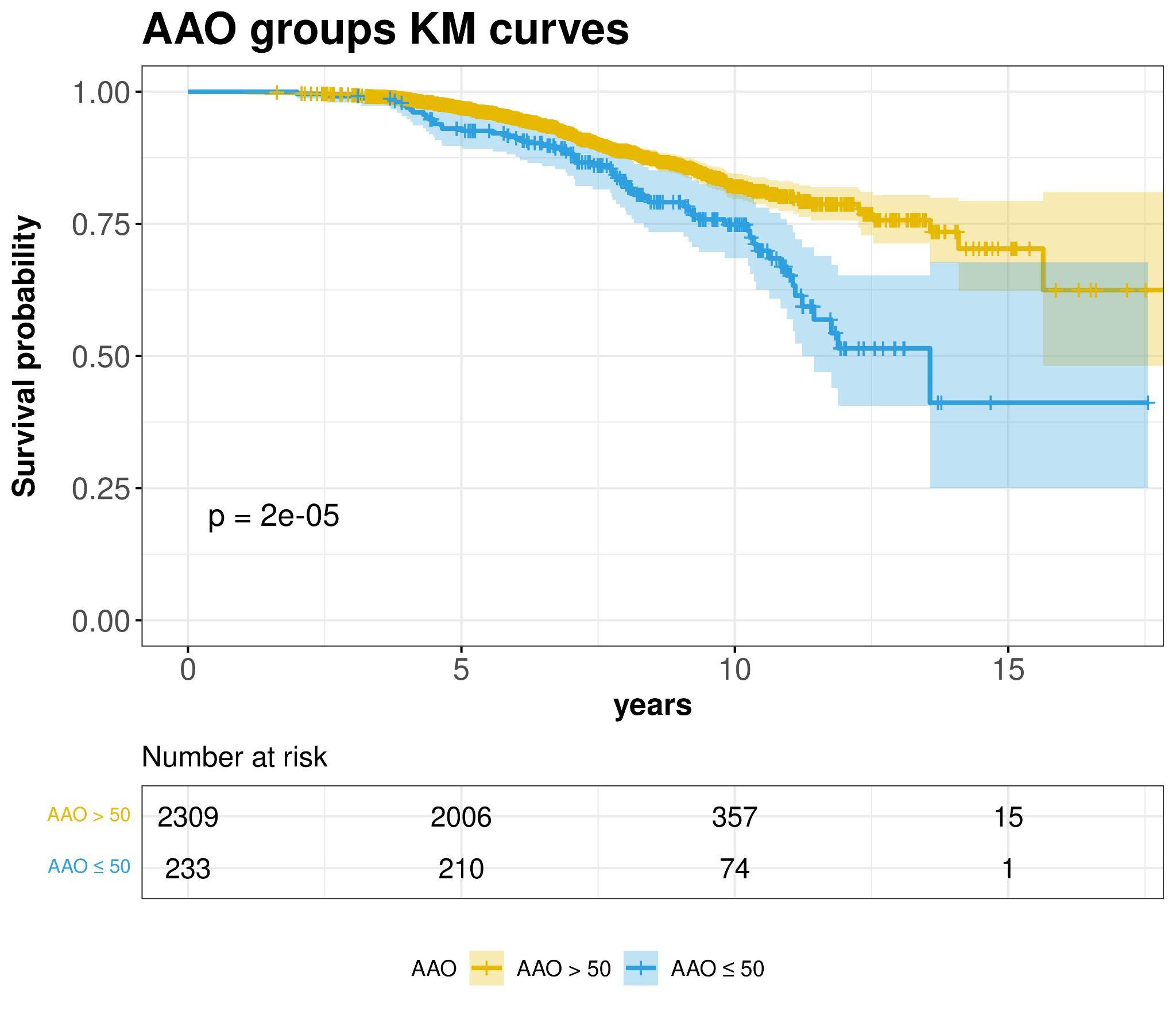

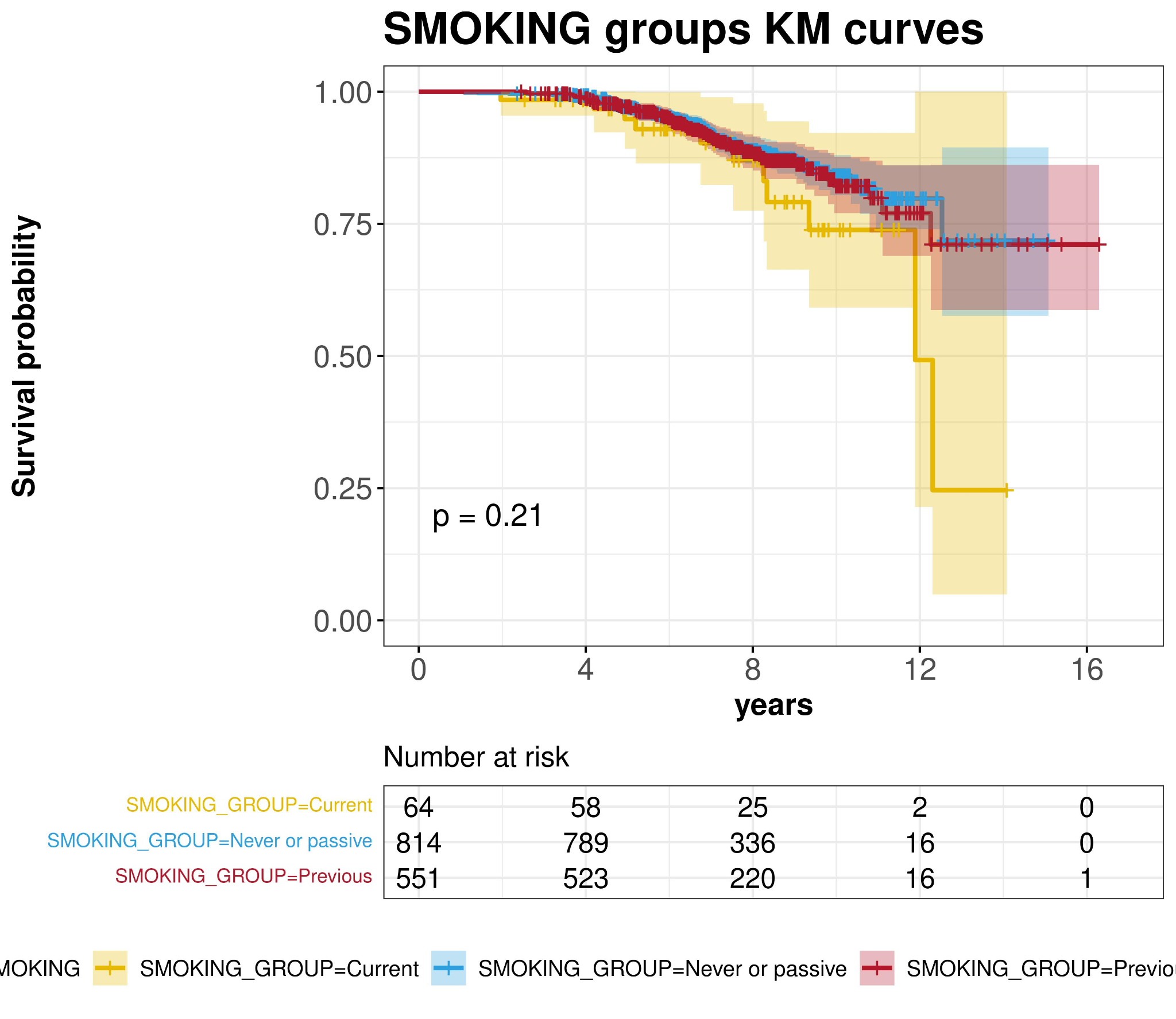

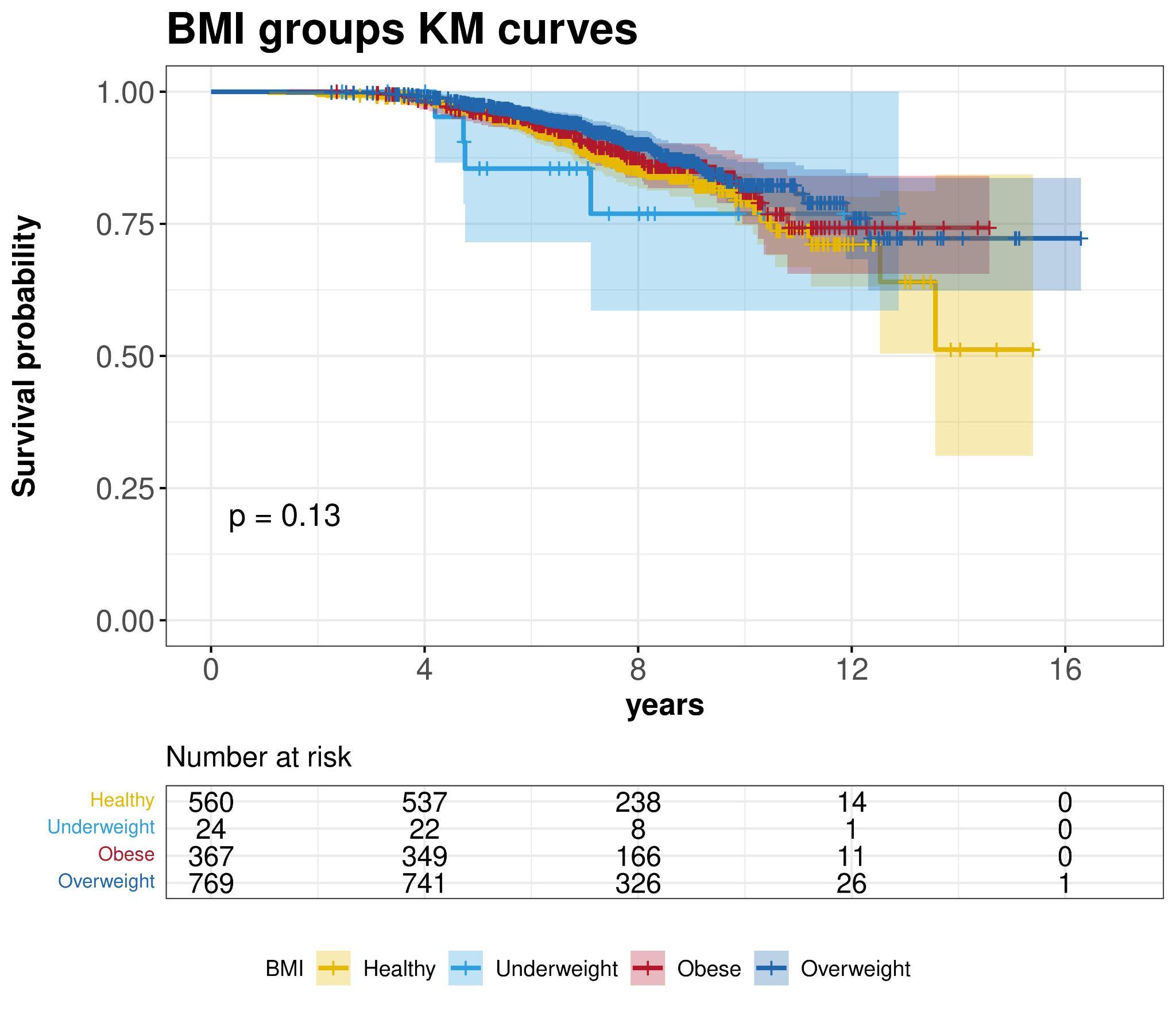

Previous = Previous smoker from baseline

Current = Current smoker at baseline

Never or passive = Never or passive smoker at baseline

**eFigure 3**. **Power calculation and simulation**. Power calculation and simulation to detect genetic association with time to develop LID as a function of sample size, relative allele frequency (AF), and genetic hazard ratio (GHR). a) Power calculation (y-axis) for the current sample size based on different AFs (x-axis) (graph label) ; b) Power simulation to explore the increase in power (y-axis) to detect lower GHR (graph grid) and relative AFs (graph label) as we increase the sample size (x-axis)

**a	b**

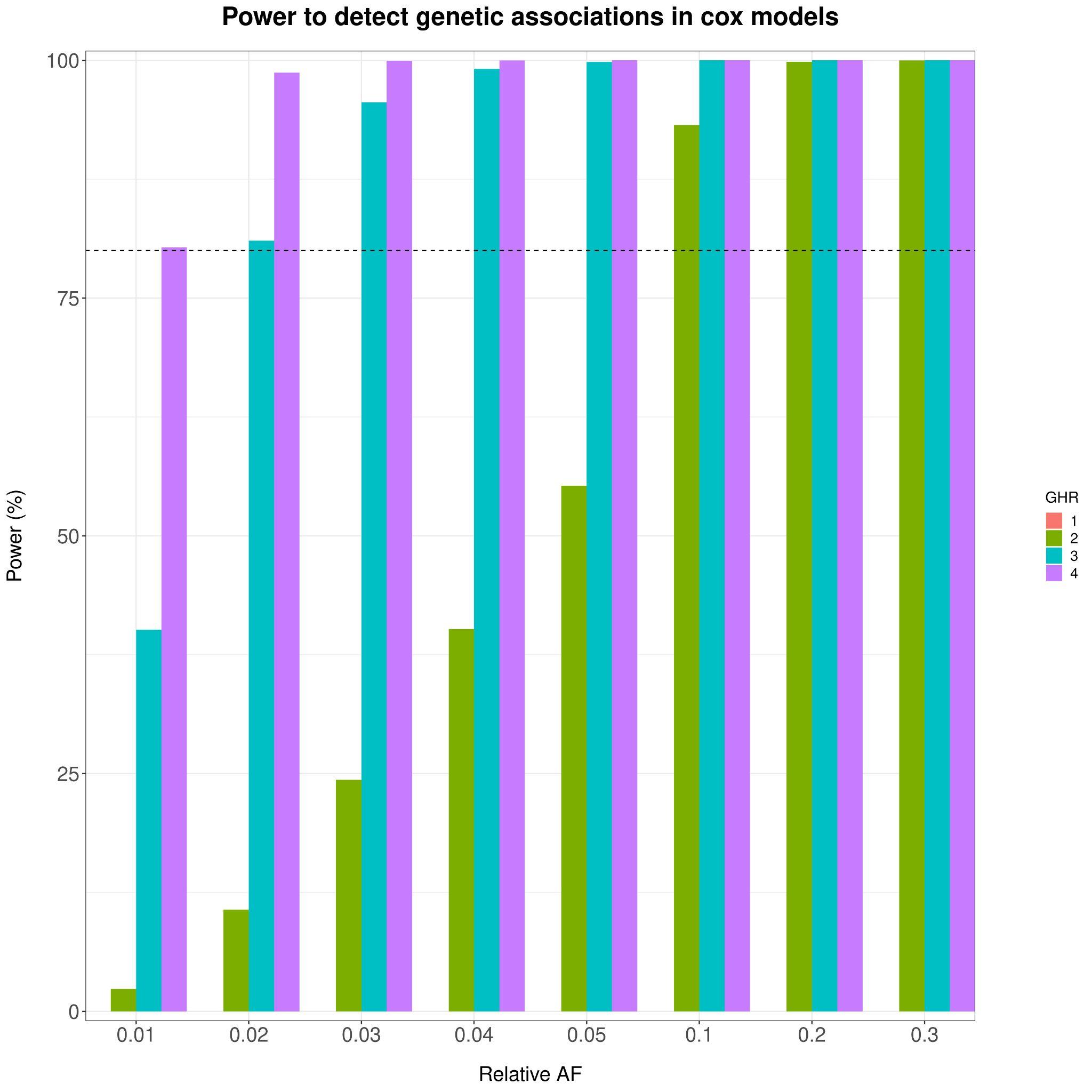

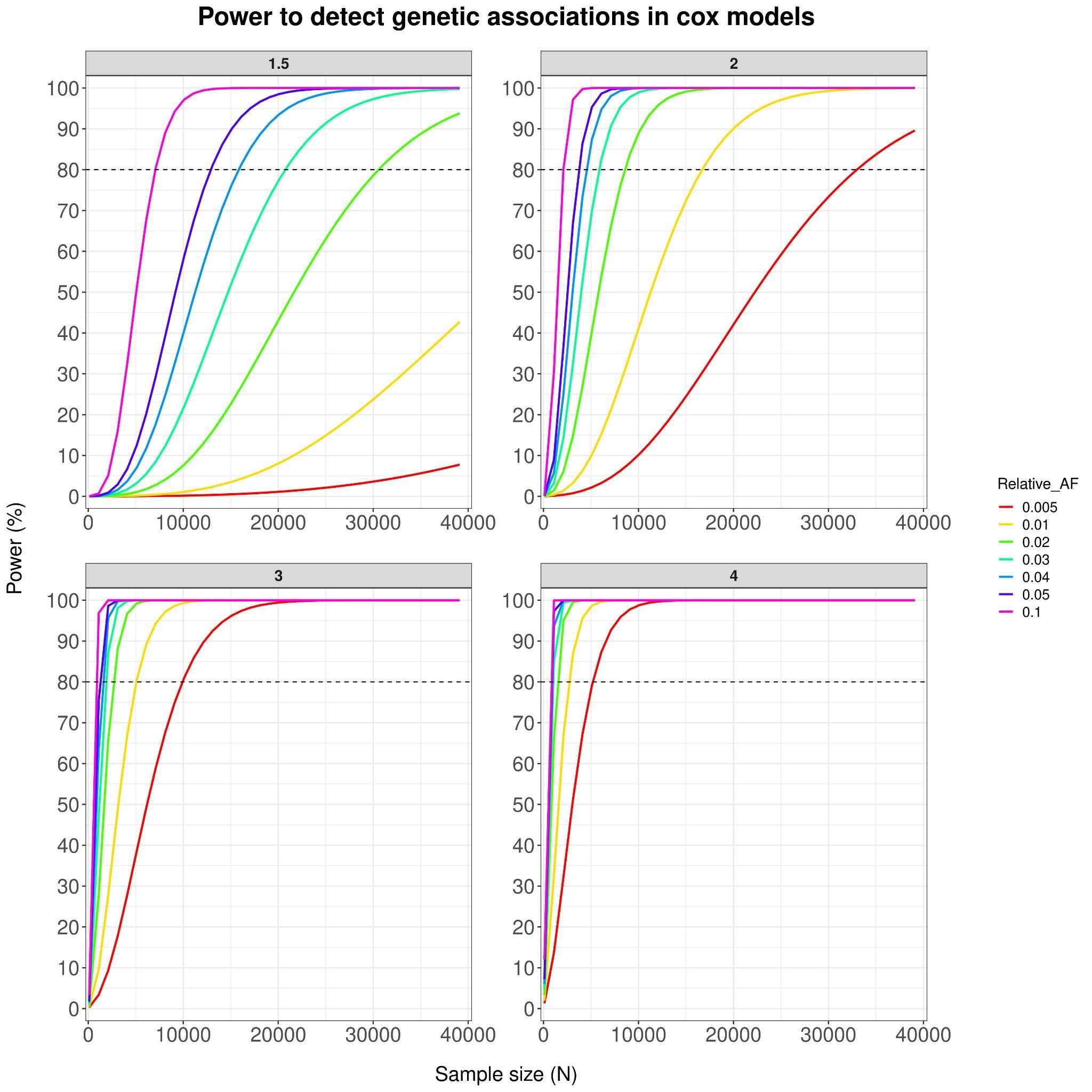

**eFigure 4. SNP metrics correlation between the CPH basic and best models.** Correlation between SNP regression coefficients (A) and SNP test-statistic (B) in from the basic and the adjusted model. The Pearson correlation coefficient ® and the significance (p) is given.

**a	b**

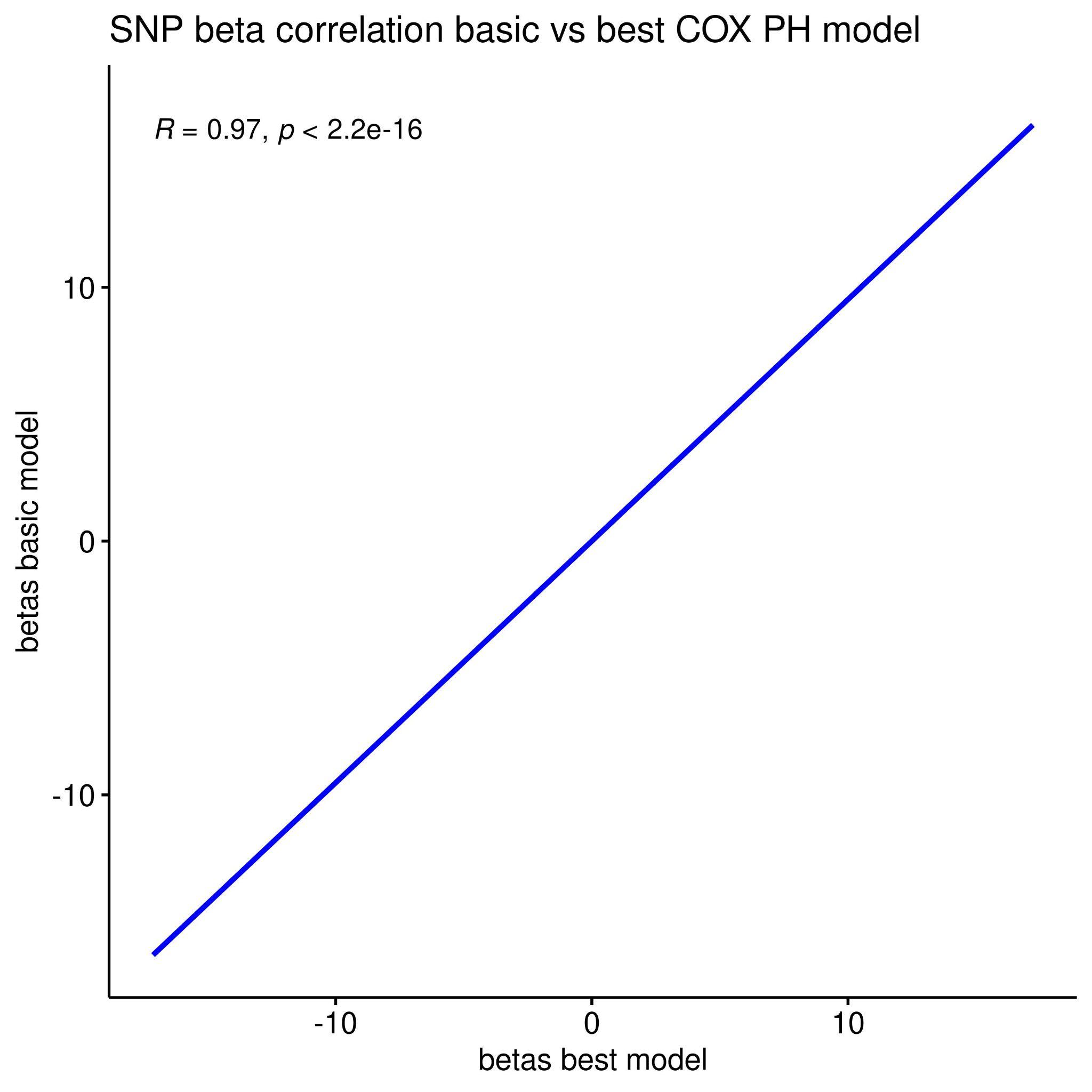

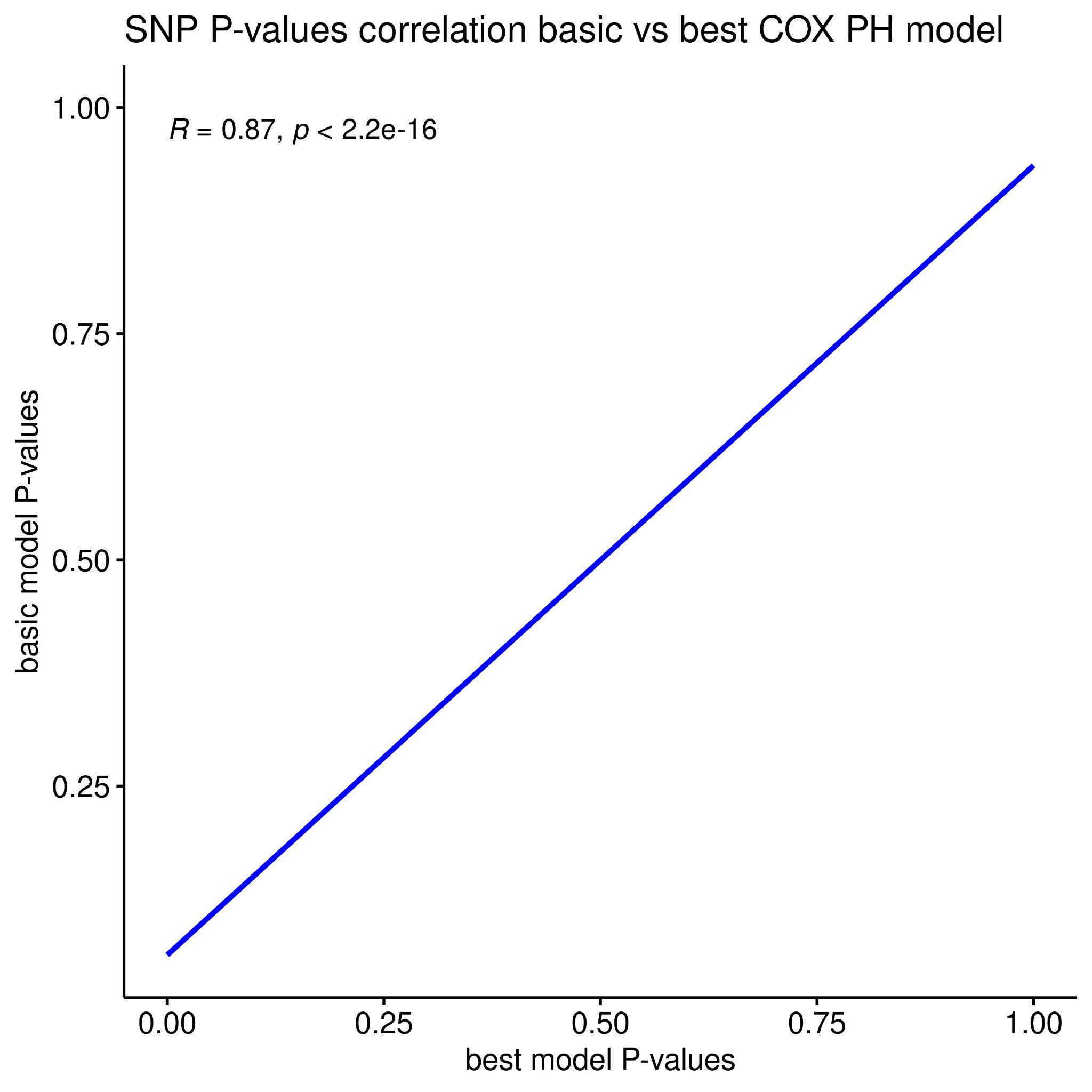

**eFigure 5.** **LRP8 locus fine-mapping and top 5 TFBS marks.** From top to bottom, *LRP8* transcript plot, locus plot, the fine-mapping nominated variants across fine-mapping tools, and the top 4 ENCODE TFBS peaks in the *XYLT1* locus. In the locus plot, the SNPs are coloured in red as LD (given by R2) increases, and blue as the LD decreases. In the fine-mapping track, we highlight the SNPs with the highest posterior probabilities for each fine-mapping tool (ABF, FINEMAP, SUSIE, POLYFUN_SUSIE). In addition, we highlight in yellow the Consensus SNP with the highest mean Posterior Probability (mean). The next four rows show the transcription factor binding sites (TFBS) densities (y-axis) measured on different cell lines and laboratories. XGR finds the top 5 transcription factors (TF) with the highest binding activity in the track genomic window. These top 5 TF are displayed in the Assay label.

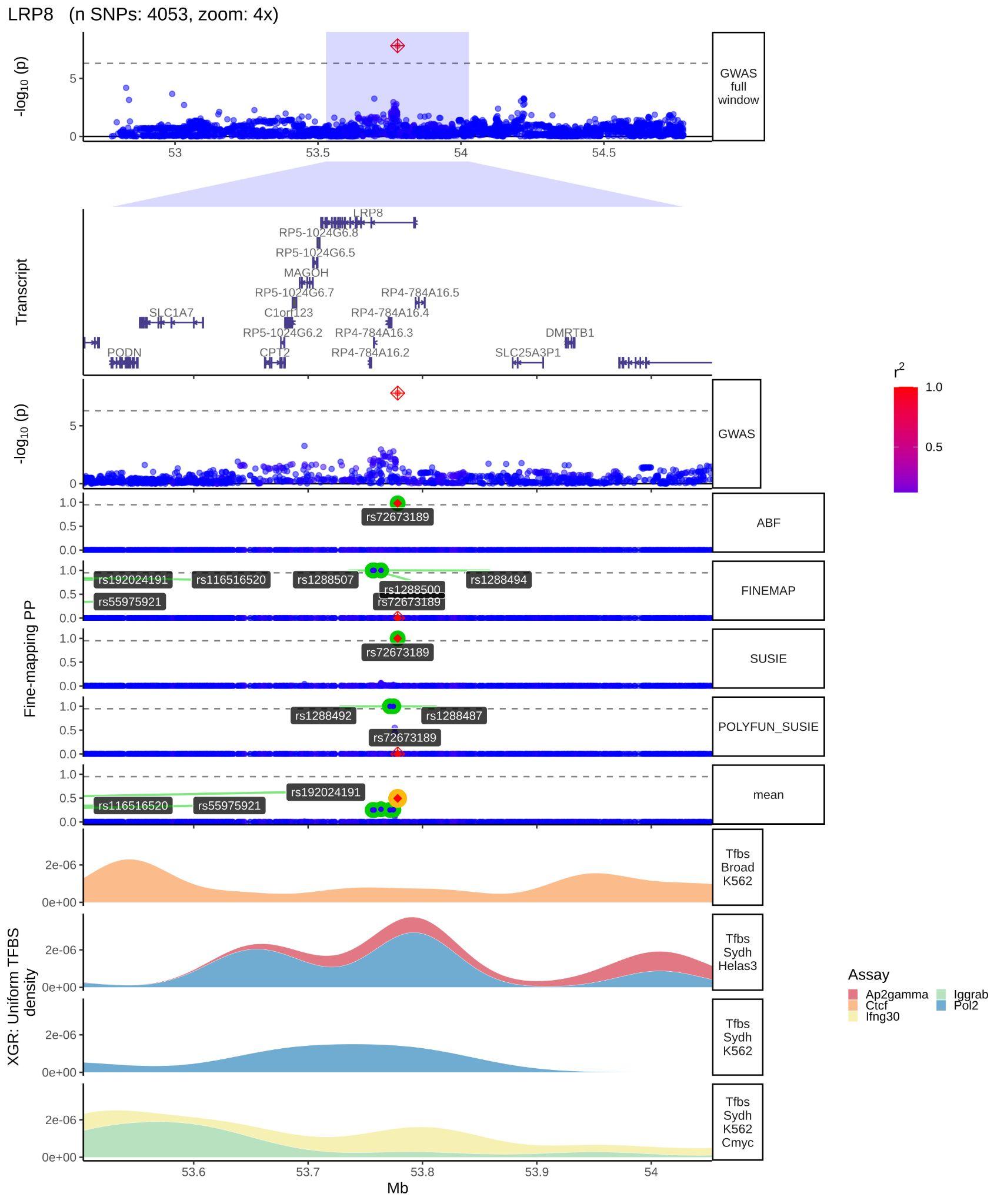

**eFigure 6. XYLT1 locus fine-mapping and brain cell type specific regulatory marks.** From top to bottom, transcript plot, locus plot, the fine-mapping nominated variants across fine-mapping tools, brain cell type specific regulatory element marks. In the locus plot, the SNPs are coloured in red as LD (given by R2) increases, and blue as the LD decreases. In the fine-mapping track, we highlight the SNPs with the highest posterior probabilities for each fine-mapping tool (ABF, FINEMAP, SUSIE, POLYFUN_SUSIE). In addition, we highlight in yellow the Consensus SNP with the highest mean Posterior Probability (mean). In the cell type specific regulatory element marks, the first 4 rows are the density marks (y-axis) from ATAC-seq assay (in pink), and CHIP-seq assays (H3K27ac in blue, and H3K4me3 in cyan), in astrocytes, microglia, neurons, and oligodendrocytes. The next four rows are the distal anchored chromatin loops (black curves). We see how, only in neurons, there is a chromatin loop forming from the *XYL71* GWS and the fine-mapped consensus variant towards the LRP8 promoter (purple).

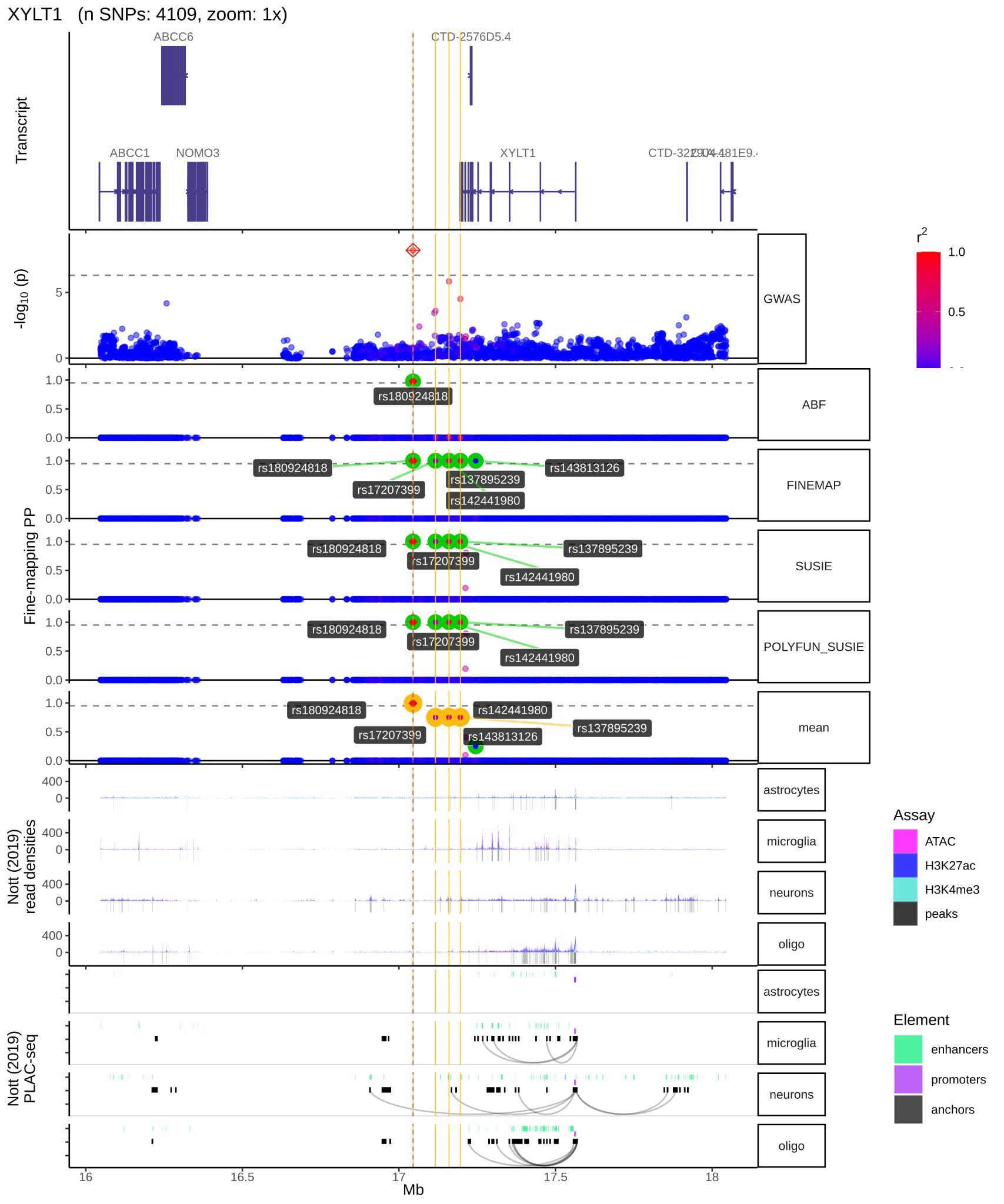

**eFigure 7.** **XYLT1 locus fine-mapping and top 5 TFBS marks.** From top to bottom, *XYLT1* transcript plot, locus plot, the fine-mapping nominated variants across fine-mapping tools, and the top 4 ENCODE TFBS peaks in the *XYLT1* locus. In the locus plot, the SNPs are coloured in red as LD (given by R2) increases, and blue as the LD decreases. In the fine-mapping track, we highlight the SNPs with the highest posterior probabilities for each fine-mapping tool (ABF, FINEMAP, SUSIE, POLYFUN_SUSIE). In addition, we highlight in yellow the Consensus SNP with the highest mean Posterior Probability (mean). The next four rows show the transcription factor binding sites (TFBS) densities (y-axis) measured on different cell lines and laboratories. XGR finds the top 5 transcription factors (TF) with the highest binding activity in the track genomic window. These top 5 TF are displayed in the Assay label.

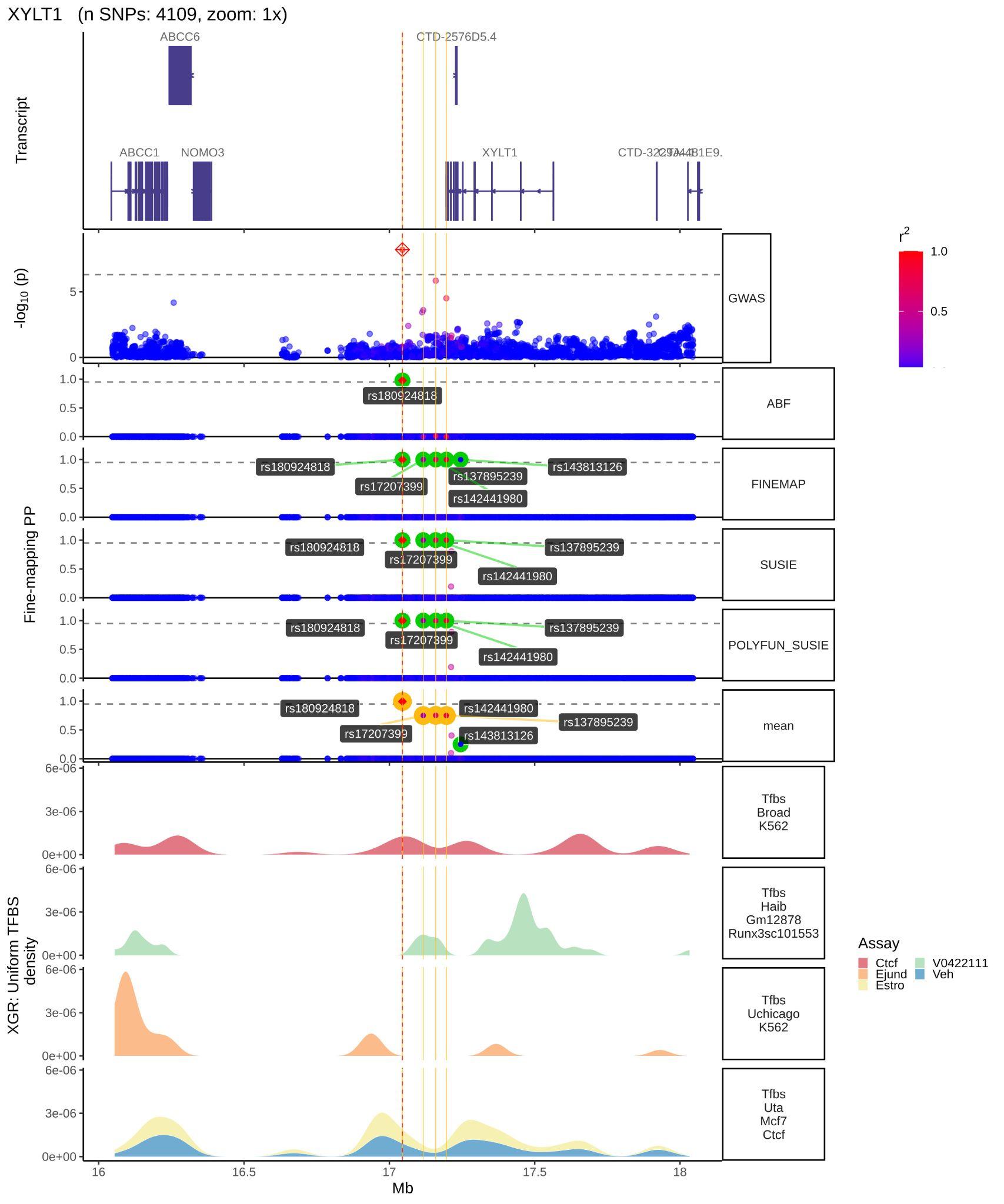

**eFigure 8.** **PRS Receiver operating characteristic (ROC) curves**. ROC curves for patient specific PRS measures derived on TPD (A) and OPDC (B) cohorts. The red curve represents the area under the curve (AUC) true positive (sensitivity) versus false positive rate ( 1 - Specificity) rate.

**a	b**

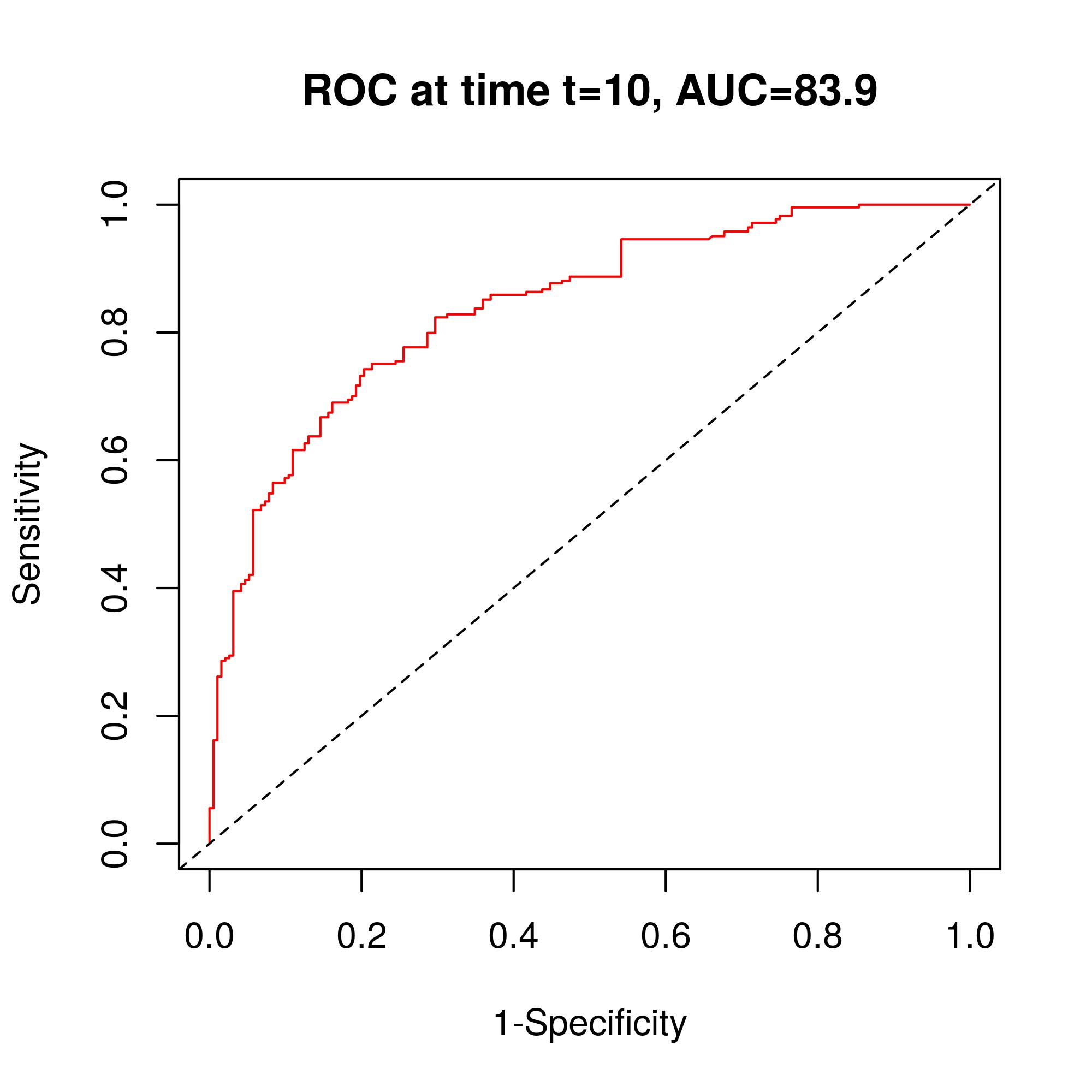

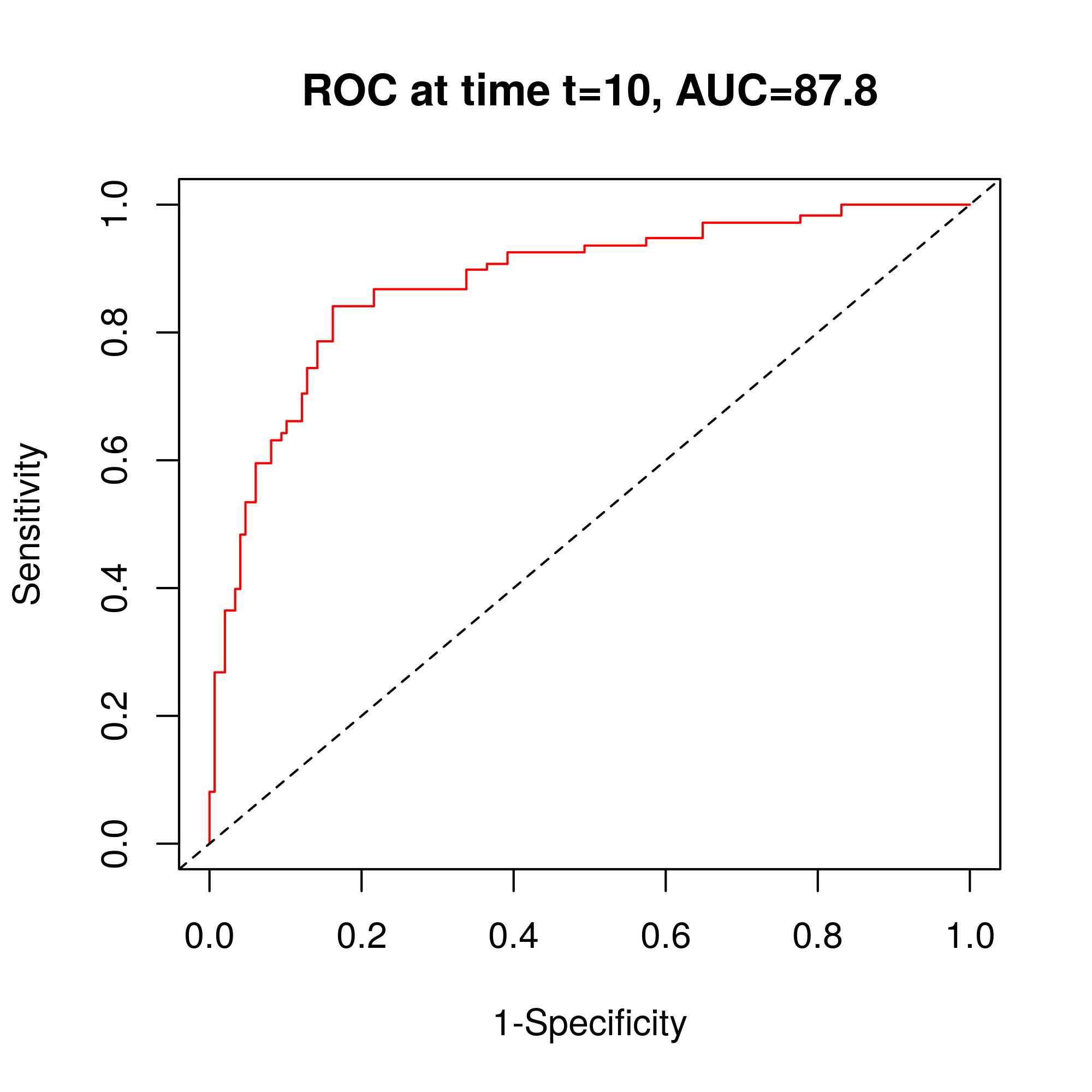

**4. eData.**

**eData 1. List of all covariates included in the stepwise regression model**

SEX,MDSUPDRSIII,PC1,PC2,PC3,PC4,PC5,AAO.std,Ldopa_previous,disease_duration_diag,Ldopa_bin,LEDD_levo,LEDD_total_V1,Amantadine_bin_V1,Anticholinergic_bin_V1,COMTI_bin_V1,DA_bin_V1,MAOBI_bin_V1,drug_naive_V1,Centre,dob,disease_duration,disease_duration_diag_cat,time_onset_to_diag,ethnicity_all,demo_QRISK,handedness,religion,marital_status,car_ownership,housing_tenure,years_education_bin,BMI,weight,height,Pulse_rate_lying,Pulse_rate_standing,Systolic_BP_lying,Systolic_BP_standing,Diastolic_BP_lying,Diastolic_BP_standing,SBP_postural_drop,DBP_postural_drop,pulse_postural_drop,orthostatic_hypotension,probability_pd,PDQ8_total,ESS_total,RBD_total,restless_legs,EQ5D_index,EQ5D_vas_score,constip_cat,gastro_total,Leeds_anx_total,Leeds_anx_bin,Leeds_dep_total,Leeds_dep_bin,QUIP_all,MOCA_total,seman_flu_score,scopa_aut_gastro,scopa_aut_urinary,scopa_aut_cardio,scopa_aut_thermo,scopa_aut_puppillo,NMSS_cardio,NMSS_sleep,NMSS_mood,NMSS_perception,NMSS_attention,NMSS_gastro,NMSS_urinary,NMSS_sexual,NMSS_misc,PDSS_total,LEDD_total,drug_naive,Amantadine_bin,Anticholinergic_bin,COMTI_bin,DA_bin,MAOBI_bin,Amantadine_previous,Anticholinergic_previous,COMTI_previous,DA_previous,MAOBI_previous,hoehn_and_yahr_stage,Schwab_england,UPDRS_I_total,UPDRS_II_total,patients_clinical_state,UPDRS_hallucinations,UPDRS_fatigue,UPDRS_pain,UPDRS_constipation,UPDRS_rigidity,UPDRS_bradykinesia,UPDRS_laterality,UPDRS_laterality_bin,side_affected_onset,FSAD_a1,FSAD_a2,FSAD_a3,FSAD_a4,FSAD_a41,FSAD_a42,FSAD_a43,FSAD_a45,FSAD_other_text,SCF_tremor_a11,SCF_tremor_a12,SCF_tremor_a13,SCF_tremor_a14,SCF_tremor_other_text,SCF_rigid_a21,SCF_rigid_a22,SCF_rigid_a23,SCF_rigid_a24,SCF_rigid_a25,SCF_rigid_a26,SCF_rigid_other_text,SCF_brady_a31,SCF_brady_a32,SCF_brady_a33,SCF_brady_a34,SCF_brady_a35,SCF_brady_other_text,SCF_posture_a41,SCF_posture_a42,SCF_posture_a43,SCF_posture_a45,SCF_posture_other_text,SCF_hyperkinesias_a51,SCF_hyperkinesias_a52,SCF_hyperkinesias_a53,SCF_hyperkinesias_a54,SCF_hyperkinesias_other_text,SCF_hemiatrophy_a6,SCF_autonomic_a71,SCF_autonomic_a73,SCF_oculomotor_a8,SCF_eyelid_a9,SCF_otherneuro_Hyperreflexia,SCF_otherneuro_Babinski_sign,SCF_otherneuro_Sensory_deficit,SCF_otherneuro_Amyotrophy,SCF_otherneuro_Limb_apraxia,SCF_otherneuro_Sleep_apnea,SCF_otherneuro_Dysmetria,SCF_otherneuro_other,SCF_response_a11,SCF_tachyphemia_a12,SCF_unusual_Presentation,SCF_unusual_Symptoms,SCF_unusual_Course,SCF_unusual_response,SCF_unusual_Other,SCF_specify_unusual,history_angina,history_heart_fail,history_stroke,history_heart_att,history_diabetes,type_1_diabetes,type_2_diabetes,history_cholesterol,history_high_bp,vascular_cat,heart_disease,vascular_risk,history_bowel,history_prostate,history_breast,history_asthma,history_bronch,history_emphysema,history_arthritis,history_other,adopted,mother_pd,father_pd,matgrand_pd,patgrand_pd,matauun_pd,patauun_pd,sum_fmhx_pd,first_degree_rel_pd,second_degree_rel_pd,recessive_PD_history,dominant_PD_history,mother_stroke,mother_dement_and_alz,father_stroke,father_dement_and_alz,brother_stroke,brother_dement_and_alz,sister_stroke,sister_dement_and_alz,halfsib_dement_and_alz,matgrandfather_stroke,matgrandfather_dement_and_alz,matgrandmother_stroke,matgrandmother_dement_and_alz,patgrandfather_stroke,patgrandfather_dement_and_alz,patgrandmother_stroke,patgrandmother_dement_and_alz,matau_stroke,matau_dement_and_alz,patau_stroke,patau_dement_and_alz,mother_only_stroke,maternal_stroke,father_only_stroke,paternal_stroke,mother_only_dement_and_alz,maternal_dement_and_alz,father_only_dement_and_alz,paternal_dement_and_alz,drugs_cardio,drugs_cardio_group_1,drugs_cardio_group_2,drugs_cardio_group_3,drugs_cardio_group_4,drugs_cardio_group_5,drugs_cardio_group_6,drugs_cardio_group_7,drugs_cardio_group_8,drugs_cardio_group_9,drugs_lipid,drugs_diabetic,drugs_diabetic_insulin,drugs_diabetic_oral,drugs_painkill,drugs_painkill_nsai,drugs_antiplate,drugs_anticoags,drugs_asthmacopd,drugs_asthmacopd_broncho,drugs_asthmacopd_corticost,drugs_laxatives,drugs_bladder,drugs_antidepress,CISIPD_Disability,CISIPD_Motor_complications,CISIPD_Cognitive_status,merqp_smoking,smoke_QRISK,coffee_prior_diag,coffee_current,merqp_a1_work_pesticide,merqp_a2_home_pesticide,merqp_a3_work_solvents,merqp_a4_work_heavymetals,merqp_a5_work_chemicals_fumes,merqp_a5_1,merqp_a12_oophorectomy,merqp_a13_depression,merqp_a14_anxiety,BFI_extra_total,BFI_agree_total,BFI_consci_total,BFI_neuro_total,BFI_open_total,CT_Atrophy,CT_Small_vessel_disease,CT_Lacunar_infarction,CT_Territory_infarction,CT_Hydrocephalus,CT_Other_abnormality,MRI_Atrophy,MRI_Small_vessel_disease,MRI_Lacunar_infarction,MRI_Territory_infarction,MRI_Hydrocephalus,MRI_Other_abnormality,FPCIT_SPECT_res,F_DOPA_PET_res,EQ5D_1,EQ5D_2,EQ5D_3,EQ5D_4,EQ5D_5,LADS_1,LADS_2,LADS_3,LADS_4,LADS_5,LADS_6,LADS_7,LADS_8,LADS_9,LADS_10,LADS_11,LADS_12,constip1_bowelfreq,constip2_laxative,constip3_fruitvegeuse,constip4_exercise,GCSI_a1,GCSI_a2,GCSI_a3,GCSI_a4,GCSI_a5,GCSI_a6,GCSI_a7,GCSI_a8,saut_a1_,saut_a2_,saut_a3_,saut_a4_,saut_a5,saut_a6_,saut_a7_,saut_a14,saut_a15,saut_a16,saut_a17,saut_a18,saut_a19,saut_a20,saut_a21,saut_a8_,saut_a9_,saut_a10,saut_a11,saut_a12,saut_a13,NMSS_severity_score_1,NMSS_severity_score_2,NMSS_severity_score_3,NMSS_severity_score_4,NMSS_severity_score_5,NMSS_severity_score_6,NMSS_severity_score_7,NMSS_severity_score_8,NMSS_severity_score_9,NMSS_severity_score_10,NMSS_severity_score_11,NMSS_severity_score_12,NMSS_severity_score_13,NMSS_severity_score_14,NMSS_severity_score_15,NMSS_severity_score_16,NMSS_severity_score_17,NMSS_severity_score_18,NMSS_severity_score_19,NMSS_severity_score_20,NMSS_severity_score_21,NMSS_severity_score_22,NMSS_severity_score_23,NMSS_severity_score_24,NMSS_severity_score_25,NMSS_severity_score_26,NMSS_severity_score_27,NMSS_severity_score_28,NMSS_severity_score_29,NMSS_severity_score_30,NMSS_frequency_score_1,NMSS_frequency_score_2,NMSS_frequency_score_3,NMSS_frequency_score_4,NMSS_frequency_score_5,NMSS_frequency_score_6,NMSS_frequency_score_7,NMSS_frequency_score_8,NMSS_frequency_score_9,NMSS_frequency_score_10,NMSS_frequency_score_11,NMSS_frequency_score_12,NMSS_frequency_score_13,NMSS_frequency_score_14,NMSS_frequency_score_15,NMSS_frequency_score_16,NMSS_frequency_score_17,NMSS_frequency_score_18,NMSS_frequency_score_19,NMSS_frequency_score_20,NMSS_frequency_score_21,NMSS_frequency_score_22,NMSS_frequency_score_23,NMSS_frequency_score_24,NMSS_frequency_score_25,NMSS_frequency_score_26,NMSS_frequency_score_27,NMSS_frequency_score_28,NMSS_frequency_score_29,NMSS_frequency_score_30,NMSS_1,NMSS_2,NMSS_3,NMSS_4,NMSS_5,NMSS_6,NMSS_7,NMSS_8,NMSS_9,NMSS_10,NMSS_11,NMSS_12,NMSS_13,NMSS_14,NMSS_15,NMSS_16,NMSS_17,NMSS_18,NMSS_19,NMSS_20,NMSS_21,NMSS_22,NMSS_23,NMSS_24,NMSS_25,NMSS_26,NMSS_27,NMSS_28,NMSS_29,NMSS_30,score_BFI_a1_,score_BFI_a6_,score_BFI_a11,score_BFI_a16,score_BFI_a21,score_BFI_a26,score_BFI_a31,score_BFI_a36,score_BFI_a2_,score_BFI_a7_,score_BFI_a12,score_BFI_a17,score_BFI_a22,score_BFI_a27,score_BFI_a32,score_BFI_a37,score_BFI_a42,score_BFI_a3_,score_BFI_a8_,score_BFI_a13,score_BFI_a18,score_BFI_a23,score_BFI_a28,score_BFI_a33,score_BFI_a38,score_BFI_a43,score_BFI_a4_,score_BFI_a9_,score_BFI_a14,score_BFI_a19,score_BFI_a24,score_BFI_a29,score_BFI_a34,score_BFI_a39,score_BFI_a5_,score_BFI_a10,score_BFI_a15,score_BFI_a20,score_BFI_a25,score_BFI_a30,score_BFI_a35,score_BFI_a40,score_BFI_a41,score_BFI_a44,Parents_related,Parents_relationship,Parents_other_relat_state,Person_related,Which_relatives,unusual_present,unusual_present_all,SCF_unusual_Presentation_all,SCF_unusual_Symptoms_all,SCF_unusual_Signs_all,SCF_unusual_Course_all,SCF_unusual_response_all,SCF_unusual_Other_all,prs_nominal
